# Symptom Persistence Despite Improvement in Cardiopulmonary Health – Insights from longitudinal CMR, CPET and lung function testing post-COVID-19

**DOI:** 10.1101/2021.08.03.21260940

**Authors:** Mark Philip Cassar, Elizabeth M. Tunnicliffe, Nayia Petousi, Adam J. Lewandowski, Cheng Xie, Masliza Mahmod, Azlan Helmy Abd Samat, Rachael A. Evans, Christopher E. Brightling, Ling-Pei Ho, Stefan K. Piechnik, Nick P. Talbot, David Holdsworth, Vanessa M. Ferreira, Stefan Neubauer, Betty Raman

## Abstract

**Background:** The longitudinal trajectories of cardiopulmonary abnormalities and symptoms following infection with coronavirus disease (COVID-19) are unclear. We sought to describe their natural history in previously hospitalised patients, compare this with controls, and assess the relationship between symptoms and cardiopulmonary impairment at 6 months post-COVID-19.

**Methods:** Fifty-eight patients and thirty matched controls underwent symptom-questionnaires, cardiac and lung magnetic resonance imaging (CMR), cardiopulmonary exercise test (CPET), and spirometry at 3 months following COVID-19. Of them, forty-six patients returned for follow-up assessments at 6 months.

**Findings:** At 2-3 months, 83% of patients had at least one cardiopulmonary symptom versus 33% of controls. Patients and controls had comparable biventricular volumes and function. Native cardiac T_1_ (marker of inflammation) and late gadolinium enhancement (LGE, marker of focal fibrosis) were increased in patients. Sixty percent of patients had lung parenchymal abnormalities on CMR and 55% had reduced peak oxygen consumption (pVO_2_) on CPET.

By 6 months, 53% of patients remained symptomatic. On CMR, indexed right ventricular (RV) end-diastolic volume (−4·3 mls/m^2^, *P*=0·005) decreased and RV ejection fraction (+3·2%, *P*=0·0003) increased. Native T_1_ and LGE improved and was comparable to controls. Lung parenchymal abnormalities and peak VO_2_, although better, were abnormal in patients versus controls. 31% had reduced pVO_2_ secondary to fatigue and submaximal tests. Cardiopulmonary symptoms in patients did not associate with CMR, lung function, or CPET measures.

**Interpretation:** In patients, cardiopulmonary abnormalities improve over time, though some measures remain abnormal relative to controls. Persistent symptoms at 6 months post-COVID-19 did not associate with objective measures of cardiopulmonary health.

**Funding:** NIHR Oxford and Oxford Health BRC, Oxford BHF CRE, UKRI and Wellcome Trust.

## Introduction

First described in December 2019,^1^ severe acute respiratory syndrome coronavirus-2 (SARS-CoV-2), a beta coronavirus, is responsible for coronavirus disease (COVID-19). Our understanding of how this virus came to invade human cell lines has rapidly evolved, as the role of Angiotensin-converting enzyme-2 receptors in facilitating viral entry into cells was elucidated.^2^ Angiotensin-converting enzyme-2 receptors are not only present in type II pneumocytes but are ubiquitously expressed by the vascular cells and other visceral organs.^3^ The effect of SARS-CoV-2 on the heart is of particular importance, as it can cause a range of abnormalities including myocardial dysfunction, inflammation, and ischemic damage via direct (cytotoxic) and indirect (dysregulated immune response, thrombo-inflammation) mechanisms.^4^ Myocardial injury is more common in moderate to severe infections and predictive of poor clinical outcomes among those admitted to hospital.^5^ A number of recent studies have highlighted the role of cardiac magnetic resonance imaging (CMR) and cardiopulmonary exercise testing (CPET) in evaluating the mechanisms and functional consequences of cardiopulmonary injury in COVID-19 survivors.^5-7^ Detailed assessments have typically been undertaken at a single time point within weeks to months after infection and do not reveal the natural history of cardiopulmonary pathology. A high burden of cardiopulmonary symptoms has also been reported and the role of contemporaneous investigations in elucidating the underlying cause for symptoms is unknown.

Previously, we undertook a holistic assessment of COVID-19 patients at 2-3 months following moderate to severe infection using symptom-based questionnaires, multiorgan magnetic resonance imaging (MRI), spirometry, and CPET.^8^ We observed a high prevalence of tissue abnormalities involving the heart (26%) and lungs (60%) on MRI, together with reduced forced expiratory volume in 1 second (FEV_1_) and forced vital capacity (FVC) and marked exercise intolerance on CPET in patients. Here, we sought to describe the time course evolution of cardiopulmonary symptoms, CMR, pulmonary function and CPET abnormalities in these patients from 2-3 months to 6 months and evaluate the relationship between symptoms and objective measures of cardiopulmonary health at 6 months.

This study was registered at ClinicalTrials.gov (NCT04510025) and approved in the United Kingdom by the North West Preston Research Ethics Committee (reference 20/NW/0235).

## Methods

### Study population

Fifty-eight patients with moderate to severe laboratory-confirmed (SARS-CoV-2 polymerase chain reaction positive) COVID-19, admitted for inpatient treatment at the Oxford University Hospitals National Health Service Foundation Trust, and 30 SARS-CoV-2 immunoglobulin negative controls, group-matched for age, sex, body mass index and risk factors (smoking, diabetes, and hypertension) from the community (recruited during the same period) were prospectively enrolled in this observational cohort study as previously described. A flow chart for recruitment is listed in the **Supplementary Material, p7**.

### Study procedures

Informed consent was obtained from all patients. Patient health questionnaires, cardiopulmonary magnetic resonance imaging, spirometry, CPET, electrocardiogram (ECG) and blood tests were undertaken in patients at 2-3 months and 6 months post-infection and at a single time point in controls. Gas transfer assessments were undertaken in patients at 6 months alone.

Disease severity was graded using the World Health Organisation ordinal scale for clinical improvement.^9^ Patients with severe illness were defined as those having a score of ≥5 (high flow oxygen, non-invasive and invasive ventilation).

An electrocardiogram (ECG) was performed for every participant and interpreted according to the Minnesota Code of Electrocardiographic Findings.^10^

Patient health questionnaire-15 (PHQ-15)^11^ was completed using an electronic data capture platform (CASTOR EDC, https://www.castoredc.com). The Medical Research Council (MRC) dyspnoea scale^12^ and Fatigue Severity Scale (FSS)^13^ were used to assess the prevalence and severity of breathlessness and fatigue, respectively (**Supplementary material, p3**).

CMR was carried out at 3 Tesla (Prisma, Siemens Healthineers, Erlangen, Germany) and included cine imaging to assess biventricular volumes, T_1_ and T_2_ mapping to assess myocardial inflammation and oedema, and post-contrast T_1_ mapping and late gadolinium enhancement (LGE) imaging to assess diffuse and focal/patchy fibrosis. Lung abnormalities were assessed using Half-Fourier-acquisition single□shot turbo spin□echo (HASTE) MRI before the administration of contrast (**Supplementary Material, p3**).

CMR studies were analysed using CVI42 5.11.4 (Circle Cardiovascular Imaging, Calgary, Canada). All cardiac images were analysed by CMR experts (BR, MC) (**Supplementary Material, p4**). Lung images were qualitatively assessed for parenchymal involvement by an expert radiologist (CX), with the extent of lung parenchymal opacities scored as 0 (0%), 1 (1-25%), 2 (26-50%), 3 (51-75%), or 4 (76-100%).^14^

Spirometry, including FVC and FEV_1_, was performed as per recommended guidance.^15^ Diffusion capacity for carbon monoxide (DL_CO_) and alveolar volume (Va) were measured using a ten-second single breath-hold technique with methane as the tracer gas, and adjusted for hemoglobin.^16^

Symptom-limited incremental CPET was undertaken using a cycle ergometer as previously described. Following two minutes of unloaded cycling, the work rate was increased to 20W, followed by a 10W/min ramp (**Supplementary Material, p5**).^17^

Blood-based testing consisted of complete blood count, biochemical analysis, coagulation testing, liver and renal function assessment, markers of cardiac injury (troponin T and N-terminal pro-brain natriuretic peptide/NT-proBNP), and measures of electrolytes, C-reactive protein (CRP), and procalcitonin.

Details on clinical symptoms, signs, vitals, and laboratory findings during admission were extracted from electronic medical records. ^18^

### Statistics

Continuous variables were described using mean and standard deviation for variables with parametric data across all groups. When non-parametric data was present in one or more groups, median and interquartile range (IQR) was used to facilitate comparison. Normality was assessed by the Shapiro-Wilk test. Group differences were evaluated using Student’s t-tests, Mann-Whitney U-tests, paired Student’s t-tests, and Wilcoxon Signed Ranks tests as appropriate. Categorical variables were reported as frequency and percentages, with group differences evaluated using the Chi-square test, Fisher’s exact test, Fisher-Freeman-Halton exact test, Stuart-Maxwell test, or McNemar test as appropriate. Spearman’s correlation coefficients were used to describe the relationship between two variables where relevant. Univariate binary logistic regression was used to determine the association of cardiopulmonary symptoms (chest pain, palpitations, syncope, dyspnoea, or dizziness) and objective measures of cardiopulmonary health. For any univariate association with a *P*-value of <0·05, we planned to undertake a multivariable logistic regression.

In a separate analysis, determinants of breathlessness were also ascertained (**Supplementary Material, p8)**. The conventional level of statistical significance of 5% was used. Statistical analyses were performed using SPSS Version 27.0 (IBM, Armonk, NY, USA).

## Results

Baseline characteristics of all patients and controls are listed in **Table 1**.^19^ Of the 58 patients recruited, 46 (79%) returned for follow-up assessments. Mean age of patients was 55±13 years. Thirty-four (59%) were men (**Table 1**). Thirteen (22%) belonged to Black (7/13) and Asian (6/13) ethnic groups. Twenty (34%) patients required non-invasive ventilation or intubation. Median duration of hospitalization was 9 days (IQR 5-17). The first assessment took place at a median interval of 2·3 months (IQR 2·1–2·5) from disease onset and second took place at 6·0 months (IQR 6·0 – 6·8).

**Table 1.** Demographics and baseline characteristics of COVID-19 patients who underwent single assessment, serial assessments (2-3 months & 6 months) and controls.

|  | COVID-19, 2-3m<br>(N=58) | COVID-19, 6m<br>(N=46) | Controls(N=30) | *P-values* |  |  |
| --- | --- | --- | --- | --- | --- | --- |
|  |  |  |  | 2-3m vs Controls | 6m vs Controls | 2-3m vs 6m |
| **General demographics** |  |  |  |  |  |  |
| Age, years | 55.4 (13.2) | 55.2 (13.3) | 53.9 (12.3) | 0.62 | 0.67 | 0.96 |
| Gender |  |  |  | 1.00<sup>a</sup> | 0.81<sup>a</sup> | 0.69<sup>a</sup> |
| Female | 24/58 (41.4%) | 17/46 (37.0%) | 12/30 (40.0%) |  |  |  |
| Male | 34/58 (58.6%) | 29/46 (63.0%) | 18/30 (60.0%) |  |  |  |
| BMI, kg/m<sup>2</sup> | 30.8 (26.2 - 36.4) | 30.6 (26.6 - 35.6) | 27.3 (23.1 - 35.1) | 0.17<sup>b</sup> | 0.19<sup>b</sup> | 0.91<sup>b</sup> |
| Black/Asian and minority ethnic groups | 13/58 (22.4%) | 10/46 (21.7%) | 1/30 (3.3 %) | 0.03<sup>c</sup> | 0.04<sup>c</sup> | 1.00<sup>a</sup> |
| Current/Ex-smoker | 20/58 (34.5%) | 17/46 (37.0%) | 7/30 (23.3%) | 0.34<sup>c</sup> | 0.31<sup>c</sup> | 0.84<sup>a</sup> |
| Type 1 Diabetes | 1/58 (1.7%) | 1/46 (2.2%) | 0/30 (0.0%) | 1.00<sup>c</sup> | 1.00<sup>c</sup> | 1.00<sup>c</sup> |
| Type 2 Diabetes | 8/58 (13.8%) | 7/46 (15.2%) | 3/30 (10.0%) | 0.74<sup>c</sup> | 0.73<sup>c</sup> | 1.00<sup>a</sup> |
| Hypertension | 22/58 (37.9%) | 17/46 (37.0%) | 9/30 (30.0%) | 0.49<sup>c</sup> | 0.62<sup>c</sup> | 1.00<sup>a</sup> |
| Coronary artery disease | 2/58 (3.4%) | 1/46 (2.2%) | 0/30 (0.0%) | 0.55<sup>c</sup> | 1.00<sup>c</sup> | 1.00<sup>c</sup> |
| Cerebrovascular Disease | 1/58 (1.7%) | 0/46 (0.0%) | 0/30 (0.0%) | 1.00<sup>c</sup> | 1.00<sup>c</sup> | 1.00<sup>c</sup> |
| Asthma | 20/58 (34.5%) | 17/46 (37.0%) | 6/30 (20.0%) | 0.22<sup>c</sup> | 0.13<sup>c</sup> | 0.84<sup>a</sup> |
| COPD | 3/58 (5.2%) | 2/46 (4.3%) | 0/30 (0.0%) | 0.55<sup>c</sup> | 0.51<sup>c</sup> | 1.00<sup>c</sup> |
| Previous cancer | 2/58 (3.4%) | 2/46 (4.3%) | 3/30 (10.0%) | 0.33<sup>c</sup> | 0.38<sup>c</sup> | 1.00<sup>c</sup> |
| Depression | 3/58 (5.2%) | 3/46 (6.5%) | 1/30 (3.3%) | 1.00<sup>c</sup> | 1.00<sup>c</sup> | 1.00<sup>c</sup> |
| **Admission details** |  |  |  |  |  |  |
| Median length of stay, days | 8.5 (5.0 - 17.0) | 9.0 (5.0 - 17.5) | .. | .. | .. | 0.85<sup>b</sup> |
| Readmitted | 10/58 (17.2%) | 9/46 (19.6%) | .. | .. | .. | 0.48<sup>a</sup> |
| Required ITU admission | 21/58 (36.2%) | 17/46 (37.0%) | .. | .. | .. | 0.55<sup>a</sup> |
| **qSOFA** |  |  |  |  |  |  |
| 0 | 17/58 (29.3%) | 15/46 (32.6%) | .. | .. | .. | 0.94<sup>d</sup> |
| 1 | 38/58 (65.5%) | 29/46 (63.0%) | .. | .. | .. |  |
| 2 | 3/58 (5.2%) | 2/46 (4.3%) | .. | .. | .. |  |
| 3 | 0/58 (0.0%) | 0/46 | .. | .. | .. |  |
| **Ordinal scale for clinical improvement (WHO)** |  |  |  |  |  |  |
| 1 | 0/58 (0.0%) | 0/46 | .. | .. | .. | 1.00<sup>d</sup> |
| 2 | 4/58 (6.9%) | 3/46 (6.5%) | .. | .. | .. |  |
| 3 | 22/58 (37.9%) | 16/46 (34.8%) | .. | .. | .. |  |
| 4 | 5/58 (8.6%) | 4/46 (8.7%) | .. | .. | .. |  |
| 5 | 15/58 (25.9%) | 12/46 (26.1%) | .. | .. | .. |  |
| 6 | 7/58 (12.1%) | 6/46 (13.0%) | .. | .. | .. |  |
| 7 | 5/58 (8.6%) | 5/46 (10.9%) | .. | .. | .. |  |
| **Signs and symptoms on admission** |  |  |  |  |  |  |
| Fever | 51/58 (87.9%) | 40/46 (87.0%) | .. | .. | .. | 0.56<sup>a</sup> |
| Malaise | 51/58 (87.9%) | 41/46 (89.1%) | .. | .. | .. | 0.55<sup>a</sup> |
| Shortness of breath | 51/58 (87.9%) | 41/46 (89.1%) | .. | .. | .. | 0.55<sup>a</sup> |
| Cough | 35/58 (60.3%) | 26/46 (56.5%) | .. | .. | .. | 0.42<sup>a</sup> |
| Dysgeusia | 29/58 (50.0%) | 21/46 (45.7%) | .. | .. | .. | 0.70<sup>a</sup> |
| Anosmia | 26/58 (44.8%) | 20/46 (43.5%) | .. | .. | .. | 1.00<sup>a</sup> |
| Diarrhoea | 17/58 (29.3%) | 13/46 (28.3%) | .. | .. | .. | 1.00<sup>a</sup> |
| Chest pain | 16/58 (27.6%) | 13/46 (28.3%) | .. | .. | .. | 1.00<sup>a</sup> |
| Headache | 13/58 (22.4%) | 12/46 (26.0%) | .. | .. | .. | 0.82<sup>a</sup> |
| Vomiting | 9/58 (15.5%) | 6/46 (13.0%) | .. | .. | .. | 0.79<sup>a</sup> |
| **Treatment** |  |  |  |  |  |  |
| Oxygen replacement | 54/58 (93.1%) | 43/46 (93.5%) | .. | .. | .. | 1.00<sup>c</sup> |
| Nasal cannula | 14/58 (24.1%) | 10/46 (21.7%) | .. | .. | .. | 1.00<sup>d</sup> |
| Simple face mask | 7/58 (12.1%) | 5/46 (10.8%) | .. | .. | .. |  |
| Venturi face mask | 6/58 (10.3%) | 5/46 (10.8%) | .. | .. | .. |  |
| High flow oxygen delivery | 7/58 (12.1%) | 5/46 (10.8%) | .. | .. | .. |  |
| CPAP | 8/58 (13.8%) | 7/46 (15.2%) | .. | .. | .. |  |
| Intubation | 12/58 (20.7%) | 11/46 (23.9%) | .. | .. | .. |  |
| ECMO | 0/58 (0%) | 0/46 | .. | .. | .. |  |
| Inotropic support | 4/58 (6.9%) | 4/46 (8.7%) | .. | .. | .. | 0.73<sup>c</sup> |
| Renal replacement therapy | 2/58 (3.4%) | 2/46 (4.3%) | .. | .. | .. | 1.00<sup>c</sup> |
| Antibiotics | 57/58 (98.3%) | 45/46 (97.8%) | .. | .. | .. | 1.00<sup>c</sup> |
| Antivirals | 4/58 (6.9%) | 2/46 (4.3%) | .. | .. | .. | 0.69<sup>c</sup> |
| Steroids | 16/58 (27.6%) | 14/46 (30.4%) | .. | .. | .. | 0.83<sup>a</sup> |
| **Acute organ injury** |  |  |  |  |  |  |
| Acute liver injury<sup>e</sup> | 18/58 (31.0%) | 18/46 (39.1%) | .. | .. | .. | 0.41<sup>a</sup> |
| Acute kidney injury<sup>f</sup> | 6/58 (10.3%) | 6/46 (13.0%) | .. | .. | .. | 0.76<sup>a</sup> |
| Acute cardiac injury<sup>g</sup> | 3/58 (5.2%) | 0/46 | .. | .. | .. | 0.25<sup>c</sup> |
| Pulmonary embolism | 7/58 (12.1%) | 6/46 (13.0%) | .. | .. | .. | 1.00<sup>a</sup> |
| Central | 1/58 (1.7%) | 0/46 | .. | .. | .. | 1.00<sup>c</sup> |
| Peripheral | 6/58 (10.3%) | 6/46 (13.0%) | .. | .. | .. | 0.76<sup>a</sup> |
| Data are mean (SD), median (IQR) and n/N (%), where N is the total number of participants with available data. P-values from independent Student's t-test, Chi-square (a), Mann-Whitney U test (b), Fisher's exact test (c), or Fisher-Freeman-Halton exact test (d), with bold values highlighting statistical significance. 2-3m = Two to three months. 6m = Six months. COPD = Chronic obstructive pulmonary disease. ITU = Intensive treatment unit. qSOFA = Quick sequential organ failure assessment. CPAP = Continuous positive airway pressure. ECMO = Extracorporeal membrane oxygenation. WHO = World health organization. e defined as blood levels of alanine aminotransferase (ALT) or aspartate aminotransferase (AST) above 3x the upper reference limit (>135 IU/L or >126 IU/L, respectively), alkaline phosphatase or gamma-glutamyltransferase above 2x the upper reference limit (>260 IU/L or >80 IU/L, respectively). f defined as an increase in serum creatinine of at least 26 umol/L within 48 hours, or 1.5 to 2-fold increase from baseline. g defined as an acute rise in hypersensitive troponin I above the 99th percentile upper reference limit (>34 ng/L). Control subjects were matched for co-morbidities as closely as possible. |  |  |  |  |  |  |

On admission, all patients had a raised CRP (>10mg/L), 47% had lymphopenia, and 21% were anaemic. By 6 months, CRP was raised in 13%, compared to none in controls (*P*=0·076), lymphocyte count normalized, and the proportion of those with anaemia was comparable to controls (11% versus 13%, *P*=1·0) (**Table 2**).

**Table 2.**
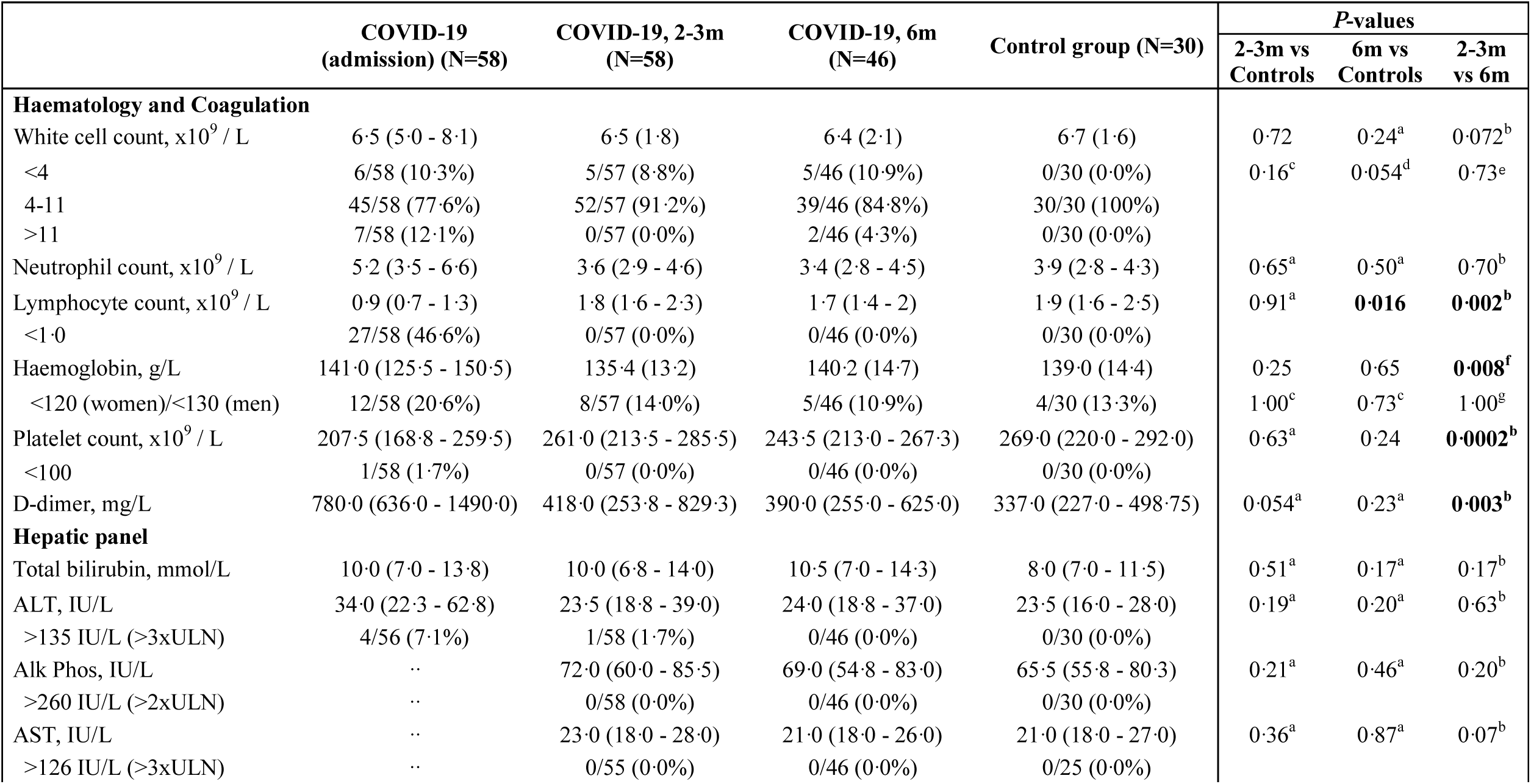
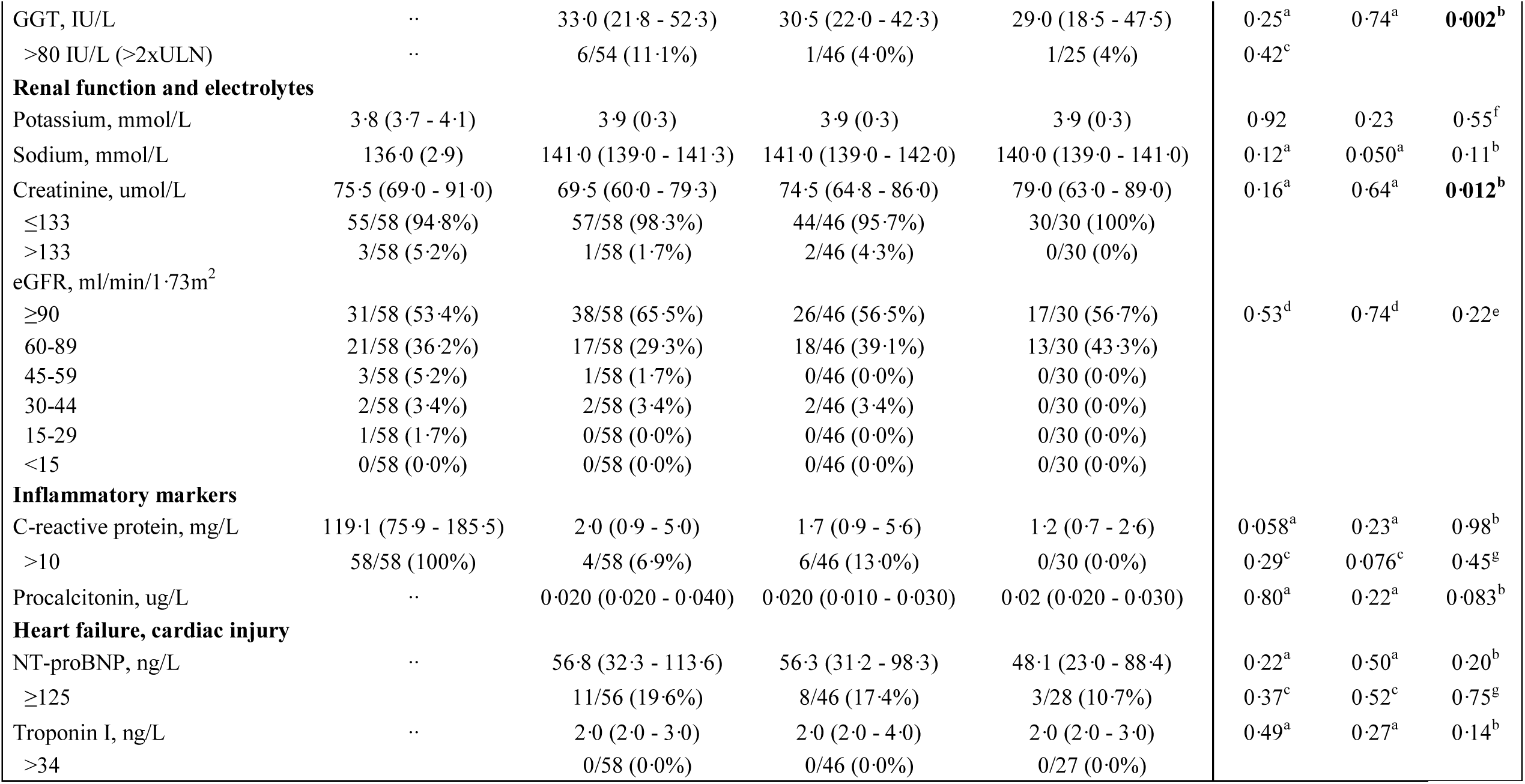
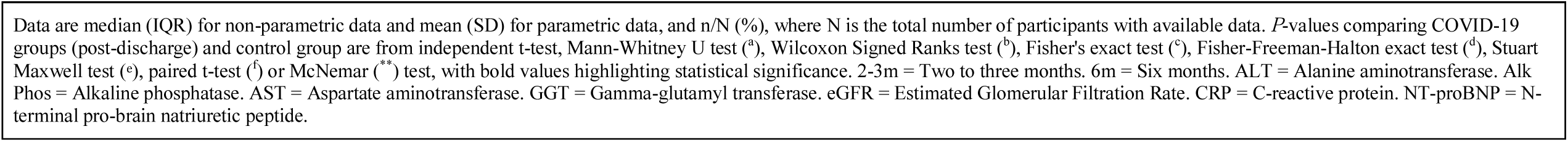
Blood test results and symptom prevalence for patients with COVID-19 and controls.

As previously reported, troponin on admission (measured in 38 patients) was abnormal in three (5%) patients. By 2-3 and 6 months, none had elevated high-sensitivity troponin levels (>34ng/L).

Only four patients had NT-proBNP measured during admission. At 2-3 months, this was elevated in 11 (20%), reducing to eight (17%) patients at 6 months versus 11% in controls (*P*=0·52).

### Electrocardiography

ECG analysis revealed atrial fibrillation in one patient at both assessments, with all other study participants (both patients and controls) demonstrating sinus rhythm.

Prevalence of bundle branch block, ST-segment elevation/depression and T wave inversion did not differ between patients (on both visits) and controls (*P*>0·05 for all variables).

### Symptom burden

Symptom prevalence in patients and controls are listed in **Table 3**. As a whole, 98% had one or more symptoms (cardiopulmonary and non-cardiopulmonary) at 2-3 months from infection, reducing to 89% by 6 months. Prevalence of cardiopulmonary symptoms (chest pain, palpitations, syncope, dyspnoea or dizziness) in patients was 83% at 2-3 months and dropped to 53% at 6 months (*P*=0·0001). At 6 months, symptoms of breathlessness (MRC) and fatigue (FSS) were worse in patients than controls (MRC grade ≥2: 57% vs 10%, *P*<0·0001; Mean FSS ≥4: 44% vs 17%, *P*=0·023, **Table 3**).

**Table 3.**
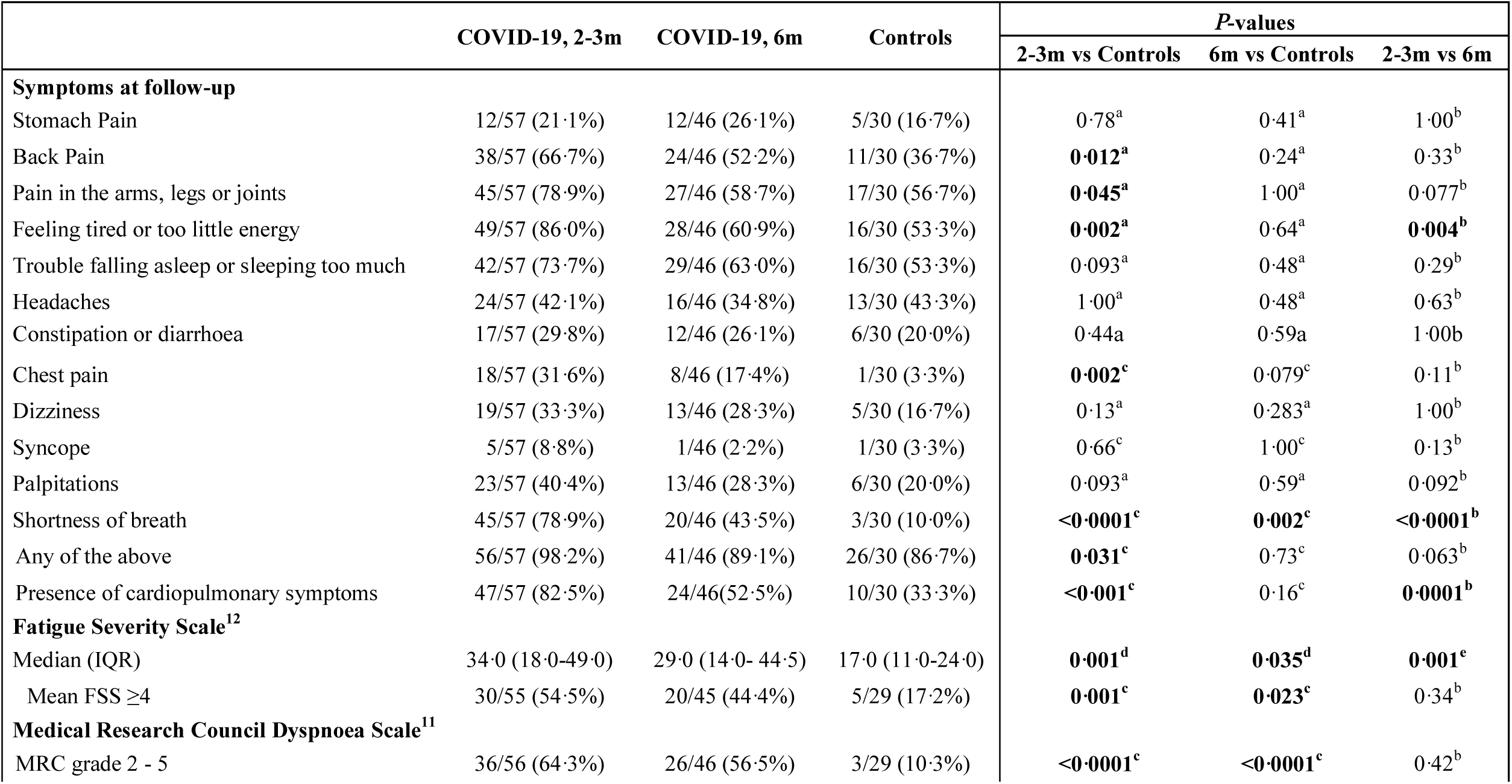
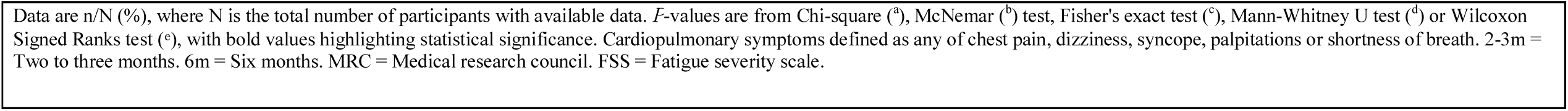
Symptom prevalence, Fatigue Severity Score and MRC dyspnoea scale in patients at follow-up and controls.

|  | COVID-19, 2-3m | COVID-19, 6m | Controls | P-values |  |  |
| --- | --- | --- | --- | --- | --- | --- |
|  |  |  |  | 2-3m vs Controls | 6m vs Controls | 2-3m vs 6m |
| Symptoms at follow-up |  |  |  |  |  |  |
| Stomach Pain | 12/57 (21.1%) | 12/46 (26.1%) | 5/30 (16.7%) | 0.78<sup>a</sup> | 0.41<sup>a</sup> | 1.00<sup>b</sup> |
| Back Pain | 38/57 (66.7%) | 24/46 (52.2%) | 11/30 (36.7%) | **0.012<sup>a</sup>** | 0.24<sup>a</sup> | 0.33<sup>b</sup> |
| Pain in the arms, legs or joints | 45/57 (78.9%) | 27/46 (58.7%) | 17/30 (56.7%) | **0.045<sup>a</sup>** | 1.00<sup>a</sup> | 0.077<sup>b</sup> |
| Feeling tired or too little energy | 49/57 (86.0%) | 28/46 (60.9%) | 16/30 (53.3%) | **0.002<sup>a</sup>** | 0.64<sup>a</sup> | **0.004<sup>b</sup>** |
| Trouble falling asleep or sleeping too much | 42/57 (73.7%) | 29/46 (63.0%) | 16/30 (53.3%) | 0.093<sup>a</sup> | 0.48<sup>a</sup> | 0.29<sup>b</sup> |
| Headaches | 24/57 (42.1%) | 16/46 (34.8%) | 13/30 (43.3%) | 1.00<sup>a</sup> | 0.48<sup>a</sup> | 0.63<sup>b</sup> |
| Constipation or diarrhoea | 17/57 (29.8%) | 12/46 (26.1%) | 6/30 (20.0%) | 0.44a | 0.59a | 1.00b |
| Chest pain | 18/57 (31.6%) | 8/46 (17.4%) | 1/30 (3.3%) | **0.002<sup>c</sup>** | 0.079<sup>c</sup> | 0.11<sup>b</sup> |
| Dizziness | 19/57 (33.3%) | 13/46 (28.3%) | 5/30 (16.7%) | 0.13<sup>a</sup> | 0.283<sup>a</sup> | 1.00<sup>b</sup> |
| Syncope | 5/57 (8.8%) | 1/46 (2.2%) | 1/30 (3.3%) | 0.66<sup>c</sup> | 1.00<sup>c</sup> | 0.13<sup>b</sup> |
| Palpitations | 23/57 (40.4%) | 13/46 (28.3%) | 6/30 (20.0%) | 0.093<sup>a</sup> | 0.59<sup>a</sup> | 0.092<sup>b</sup> |
| Shortness of breath | 45/57 (78.9%) | 20/46 (43.5%) | 3/30 (10.0%) | **<0.0001<sup>c</sup>** | **0.002<sup>c</sup>** | **<0.0001<sup>b</sup>** |
| Any of the above | 56/57 (98.2%) | 41/46 (89.1%) | 26/30 (86.7%) | **0.031<sup>c</sup>** | 0.73<sup>c</sup> | 0.063<sup>b</sup> |
| Presence of cardiopulmonary symptoms | 47/57 (82.5%) | 24/46(52.5%) | 10/30 (33.3%) | **<0.001<sup>c</sup>** | 0.16<sup>c</sup> | **0.0001<sup>b</sup>** |
| Fatigue Severity Scale<sup>12</sup> |  |  |  |  |  |  |
| Median (IQR) | 34.0 (18.0-49.0) | 29.0 (14.0- 44.5) | 17.0 (11.0-24.0) | **0.001<sup>d</sup>** | **0.035<sup>d</sup>** | **0.001<sup>e</sup>** |
| Mean FSS ≥4 | 30/55 (54.5%) | 20/45 (44.4%) | 5/29 (17.2%) | **0.001<sup>c</sup>** | **0.023<sup>c</sup>** | 0.34<sup>b</sup> |
| Medical Research Council Dyspnoea Scale<sup>11</sup> |  |  |  |  |  |  |
| MRC grade 2 - 5 | 36/56 (64.3%) | 26/46 (56.5%) | 3/29 (10.3%) | **<0.0001<sup>c</sup>** | **<0.0001<sup>c</sup>** | 0.42<sup>b</sup> |

Data are n/N (%), where N is the total number of participants with available data. *P*-values are from Chi-square (<sup>a</sup>), McNemar (<sup>b</sup>) test, Fisher's exact test (<sup>c</sup>), Mann-Whitney U test (<sup>d</sup>) or Wilcoxon Signed Ranks test (<sup>e</sup>), with bold values highlighting statistical significance. Cardiopulmonary symptoms defined as any of chest pain, dizziness, syncope, palpitations or shortness of breath. 2-3m = Two to three months. 6m = Six months. MRC = Medical research council. FSS = Fatigue severity scale.

### Serial Cardiac Imaging

Left ventricular (LV) volumes, mass, and function were not different between patients (at 2-3 months and 6 months) and controls (**Table 4**). At 6 months, two (4·5%) patients had an LV ejection fraction (LVEF) just below the cut-off of 50% (49·6 and 49·8%). Those with severe illness had lower LVEF at 6 months than other patients (60·8±6·6% vs 64·8±6·5%, *P*=0·049). None of the patients had a history of pre-existing cardiac failure.

**Table 4.**
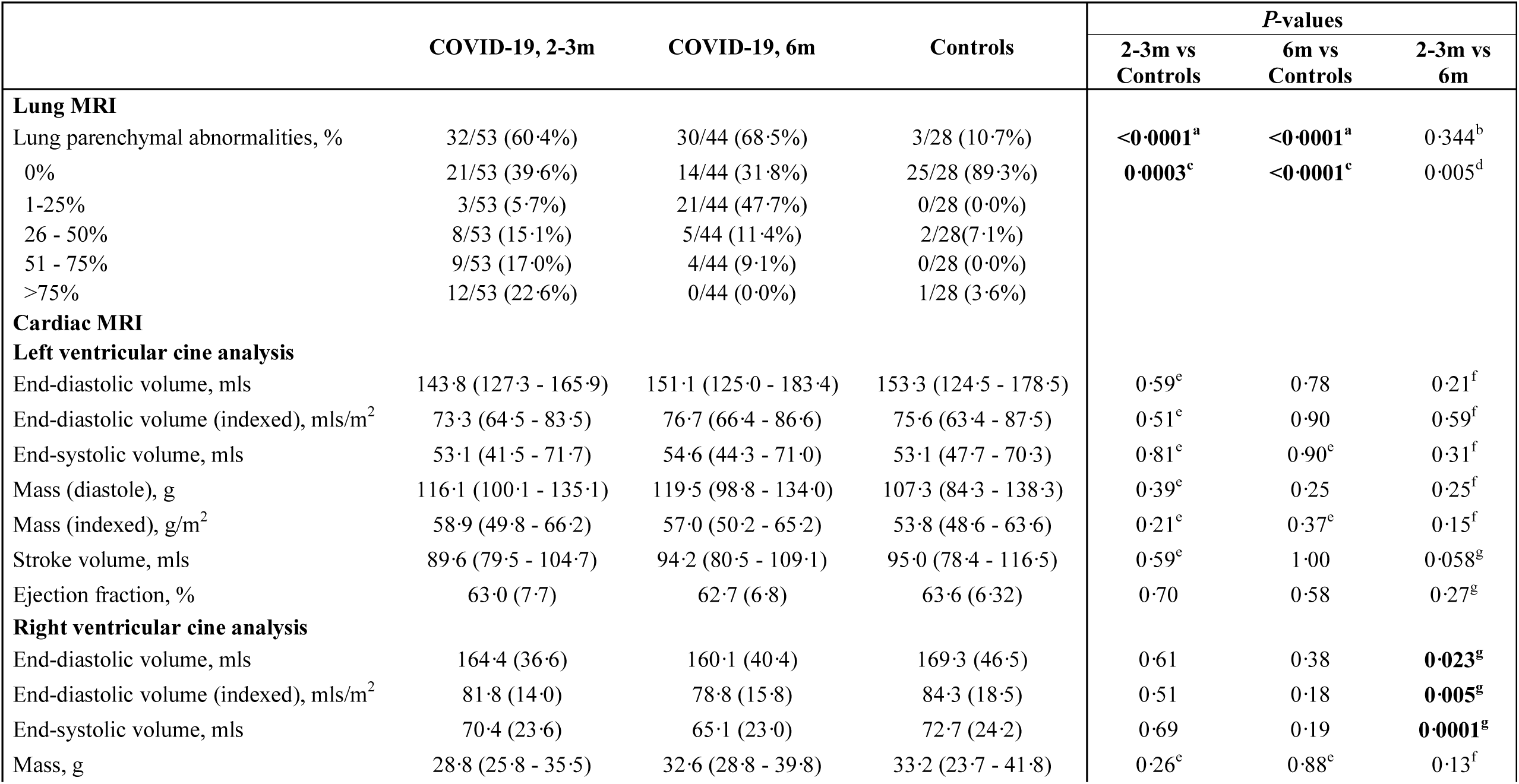
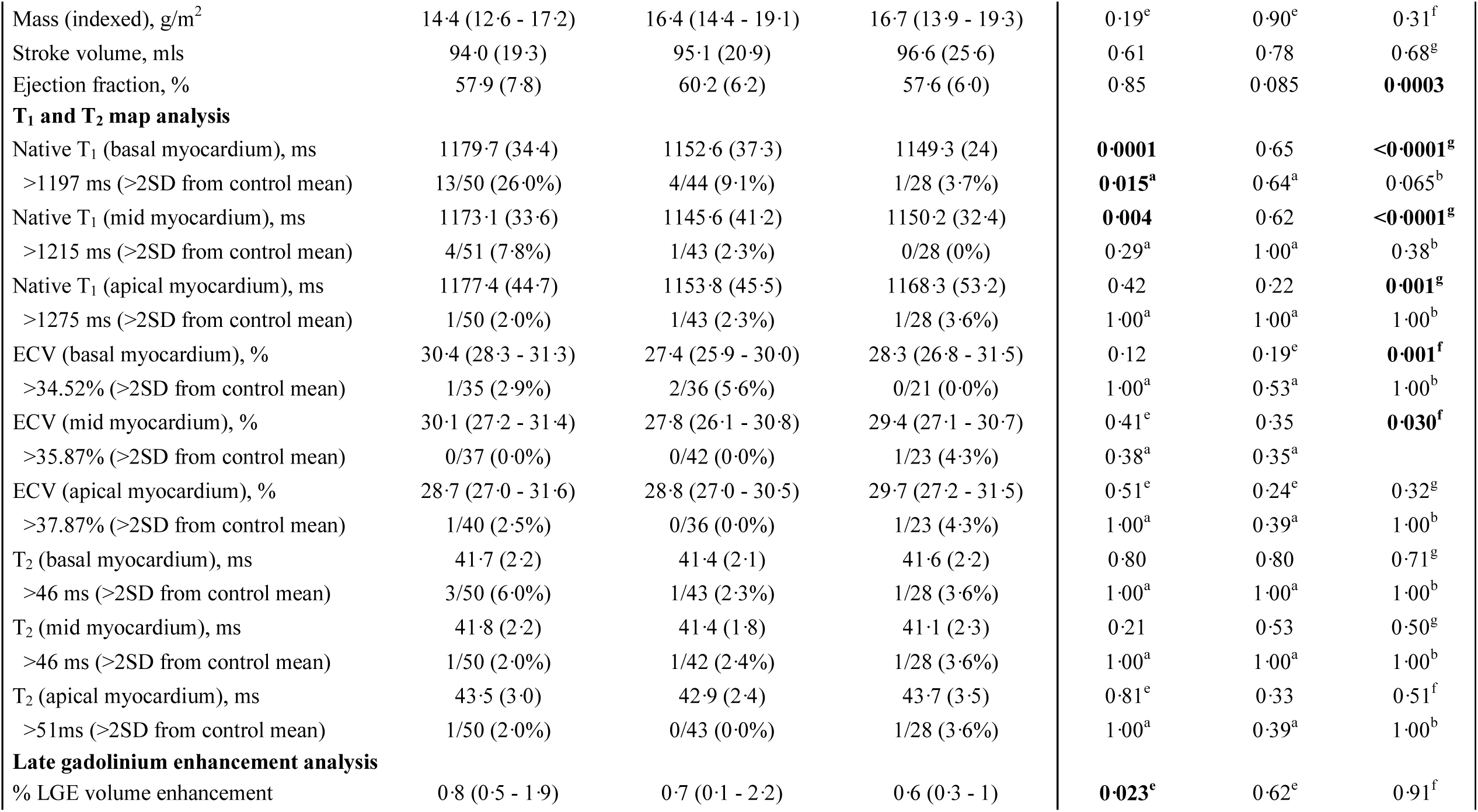
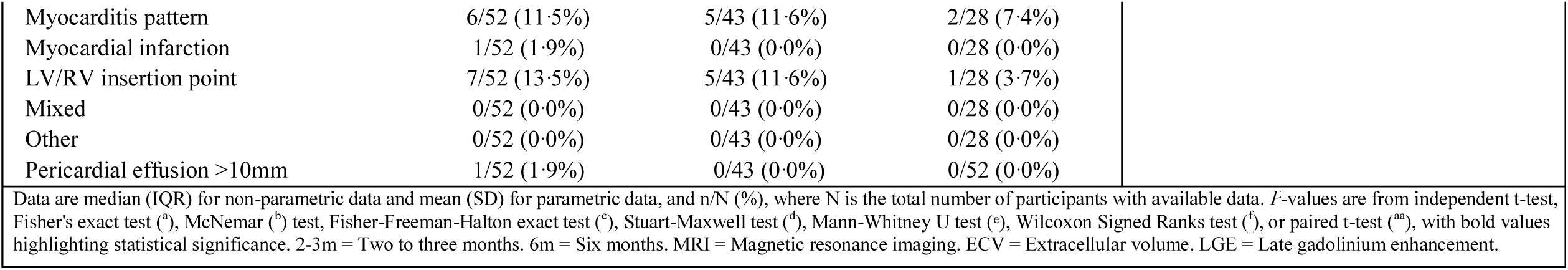
Cardiopulmonary MRI parameters in patients and controls.

|  | COVID-19, 2-3m | COVID-19, 6m | Controls | P-values |  |  |
| --- | --- | --- | --- | --- | --- | --- |
|  |  |  |  | 2-3m vs Controls | 6m vs Controls | 2-3m vs 6m |
| Lung MRI |  |  |  |  |  |  |
| Lung parenchymal abnormalities, % | 32/53 (60.4%) | 30/44 (68.5%) | 3/28 (10.7%) | <0.0001<sup>a</sup> | <0.0001<sup>a</sup> | 0.344<sup>b</sup> |
| 0% | 21/53 (39.6%) | 14/44 (31.8%) | 25/28 (89.3%) | 0.0003<sup>c</sup> | <0.0001<sup>c</sup> | 0.005<sup>d</sup> |
| 1-25% | 3/53 (5.7%) | 21/44 (47.7%) | 0/28 (0.0%) |  |  |  |
| 26 - 50% | 8/53 (15.1%) | 5/44 (11.4%) | 2/28(7.1%) |  |  |  |
| 51 - 75% | 9/53 (17.0%) | 4/44 (9.1%) | 0/28 (0.0%) |  |  |  |
| >75% | 12/53 (22.6%) | 0/44 (0.0%) | 1/28 (3.6%) |  |  |  |
| Cardiac MRI |  |  |  |  |  |  |
| Left ventricular cine analysis |  |  |  |  |  |  |
| End-diastolic volume, mls | 143.8 (127.3 - 165.9) | 151.1 (125.0 - 183.4) | 153.3 (124.5 - 178.5) | 0.59<sup>e</sup> | 0.78 | 0.21<sup>f</sup> |
| End-diastolic volume (indexed), mls/m<sup>2</sup> | 73.3 (64.5 - 83.5) | 76.7 (66.4 - 86.6) | 75.6 (63.4 - 87.5) | 0.51<sup>e</sup> | 0.90 | 0.59<sup>f</sup> |
| End-systolic volume, mls | 53.1 (41.5 - 71.7) | 54.6 (44.3 - 71.0) | 53.1 (47.7 - 70.3) | 0.81<sup>e</sup> | 0.90<sup>e</sup> | 0.31<sup>f</sup> |
| Mass (diastole), g | 116.1 (100.1 - 135.1) | 119.5 (98.8 - 134.0) | 107.3 (84.3 - 138.3) | 0.39<sup>e</sup> | 0.25 | 0.25<sup>f</sup> |
| Mass (indexed), g/m<sup>2</sup> | 58.9 (49.8 - 66.2) | 57.0 (50.2 - 65.2) | 53.8 (48.6 - 63.6) | 0.21<sup>e</sup> | 0.37<sup>e</sup> | 0.15<sup>f</sup> |
| Stroke volume, mls | 89.6 (79.5 - 104.7) | 94.2 (80.5 - 109.1) | 95.0 (78.4 - 116.5) | 0.59<sup>e</sup> | 1.00 | 0.058<sup>g</sup> |
| Ejection fraction, % | 63.0 (7.7) | 62.7 (6.8) | 63.6 (6.32) | 0.70 | 0.58 | 0.27<sup>g</sup> |
| Right ventricular cine analysis |  |  |  |  |  |  |
| End-diastolic volume, mls | 164.4 (36.6) | 160.1 (40.4) | 169.3 (46.5) | 0.61 | 0.38 | 0.023<sup>g</sup> |
| End-diastolic volume (indexed), mls/m<sup>2</sup> | 81.8 (14.0) | 78.8 (15.8) | 84.3 (18.5) | 0.51 | 0.18 | 0.005<sup>g</sup> |
| End-systolic volume, mls | 70.4 (23.6) | 65.1 (23.0) | 72.7 (24.2) | 0.69 | 0.19 | 0.0001<sup>g</sup> |
| Mass, g | 28.8 (25.8 - 35.5) | 32.6 (28.8 - 39.8) | 33.2 (23.7 - 41.8) | 0.26<sup>e</sup> | 0.88<sup>e</sup> | 0.13<sup>f</sup> |
| Mass (indexed), g/m<sup>2</sup> | 14.4 (12.6 - 17.2) | 16.4 (14.4 - 19.1) | 16.7 (13.9 - 19.3) | 0.19<sup>e</sup> | 0.90<sup>e</sup> | 0.31<sup>f</sup> |
| Stroke volume, mls | 94.0 (19.3) | 95.1 (20.9) | 96.6 (25.6) | 0.61 | 0.78 | 0.68<sup>g</sup> |
| Ejection fraction, % | 57.9 (7.8) | 60.2 (6.2) | 57.6 (6.0) | 0.85 | 0.085 | **0.0003** |
| **T<sub>1</sub> and T<sub>2</sub> map analysis** |  |  |  |  |  |  |
| Native T<sub>1</sub> (basal myocardium), ms | 1179.7 (34.4) | 1152.6 (37.3) | 1149.3 (24) | **0.0001** | 0.65 | **<0.0001<sup>g</sup>** |
| >1197 ms (>2SD from control mean) | 13/50 (26.0%) | 4/44 (9.1%) | 1/28 (3.7%) | **0.015<sup>a</sup>** | 0.64<sup>a</sup> | 0.065<sup>b</sup> |
| Native T<sub>1</sub> (mid myocardium), ms | 1173.1 (33.6) | 1145.6 (41.2) | 1150.2 (32.4) | **0.004** | 0.62 | **<0.0001<sup>g</sup>** |
| >1215 ms (>2SD from control mean) | 4/51 (7.8%) | 1/43 (2.3%) | 0/28 (0%) | 0.29<sup>a</sup> | 1.00<sup>a</sup> | 0.38<sup>b</sup> |
| Native T<sub>1</sub> (apical myocardium), ms | 1177.4 (44.7) | 1153.8 (45.5) | 1168.3 (53.2) | 0.42 | 0.22 | **0.001<sup>g</sup>** |
| >1275 ms (>2SD from control mean) | 1/50 (2.0%) | 1/43 (2.3%) | 1/28 (3.6%) | 1.00<sup>a</sup> | 1.00<sup>a</sup> | 1.00<sup>b</sup> |
| ECV (basal myocardium), % | 30.4 (28.3 - 31.3) | 27.4 (25.9 - 30.0) | 28.3 (26.8 - 31.5) | 0.12 | 0.19<sup>e</sup> | **0.001<sup>f</sup>** |
| >34.52% (>2SD from control mean) | 1/35 (2.9%) | 2/36 (5.6%) | 0/21 (0.0%) | 1.00<sup>a</sup> | 0.53<sup>a</sup> | 1.00<sup>b</sup> |
| ECV (mid myocardium), % | 30.1 (27.2 - 31.4) | 27.8 (26.1 - 30.8) | 29.4 (27.1 - 30.7) | 0.41<sup>e</sup> | 0.35 | **0.030<sup>f</sup>** |
| >35.87% (>2SD from control mean) | 0/37 (0.0%) | 0/42 (0.0%) | 1/23 (4.3%) | 0.38<sup>a</sup> | 0.35<sup>a</sup> |  |
| ECV (apical myocardium), % | 28.7 (27.0 - 31.6) | 28.8 (27.0 - 30.5) | 29.7 (27.2 - 31.5) | 0.51<sup>e</sup> | 0.24<sup>e</sup> | 0.32<sup>g</sup> |
| >37.87% (>2SD from control mean) | 1/40 (2.5%) | 0/36 (0.0%) | 1/23 (4.3%) | 1.00<sup>a</sup> | 0.39<sup>a</sup> | 1.00<sup>b</sup> |
| T<sub>2</sub> (basal myocardium), ms | 41.7 (2.2) | 41.4 (2.1) | 41.6 (2.2) | 0.80 | 0.80 | 0.71<sup>g</sup> |
| >46 ms (>2SD from control mean) | 3/50 (6.0%) | 1/43 (2.3%) | 1/28 (3.6%) | 1.00<sup>a</sup> | 1.00<sup>a</sup> | 1.00<sup>b</sup> |
| T<sub>2</sub> (mid myocardium), ms | 41.8 (2.2) | 41.4 (1.8) | 41.1 (2.3) | 0.21 | 0.53 | 0.50<sup>g</sup> |
| >46 ms (>2SD from control mean) | 1/50 (2.0%) | 1/42 (2.4%) | 1/28 (3.6%) | 1.00<sup>a</sup> | 1.00<sup>a</sup> | 1.00<sup>b</sup> |
| T<sub>2</sub> (apical myocardium), ms | 43.5 (3.0) | 42.9 (2.4) | 43.7 (3.5) | 0.81<sup>e</sup> | 0.33 | 0.51<sup>f</sup> |
| >51ms (>2SD from control mean) | 1/50 (2.0%) | 0/43 (0.0%) | 1/28 (3.6%) | 1.00<sup>a</sup> | 0.39<sup>a</sup> | 1.00<sup>b</sup> |
| **Late gadolinium enhancement analysis** |  |  |  |  |  |  |
| % LGE volume enhancement | 0.8 (0.5 - 1.9) | 0.7 (0.1 - 2.2) | 0.6 (0.3 - 1) | **0.023<sup>e</sup>** | 0.62<sup>e</sup> | 0.91<sup>f</sup> |

|  |  |  |  |
| --- | --- | --- | --- |
| Myocarditis pattern | 6/52 (11.5%) | 5/43 (11.6%) | 2/28 (7.4%) |
| Myocardial infarction | 1/52 (1.9%) | 0/43 (0.0%) | 0/28 (0.0%) |
| LV/RV insertion point | 7/52 (13.5%) | 5/43 (11.6%) | 1/28 (3.7%) |
| Mixed | 0/52 (0.0%) | 0/43 (0.0%) | 0/28 (0.0%) |
| Other | 0/52 (0.0%) | 0/43 (0.0%) | 0/28 (0.0%) |
| Pericardial effusion >10mm | 1/52 (1.9%) | 0/43 (0.0%) | 0/52 (0.0%) |
| Data are median (IQR) for non-parametric data and mean (SD) for parametric data, and n/N (%), where N is the total number of participants with available data. *P*-values are from independent t-test, Fisher's exact test <sup>(a)</sup>, McNemar <sup>(b)</sup> test, Fisher-Freeman-Halton exact test <sup>(c)</sup>, Stuart-Maxwell test <sup>(d)</sup>, Mann-Whitney U test <sup>(e)</sup>, Wilcoxon Signed Ranks test <sup>(f)</sup>, or paired t-test <sup>(aa)</sup>, with bold values highlighting statistical significance. 2-3m = Two to three months. 6m = Six months. MRI = Magnetic resonance imaging. ECV = Extracellular volume. LGE = Late gadolinium enhancement. |  |  |  |

Right ventricular (RV) volumes, mass and function did not differ between patients (at 2-3 months and 6 months) and controls (**Table 4**). In patients, indexed RV end-diastolic volume decreased (mean difference -4·3 mls/m^2^, *P*=0·005) and function (RVEF) increased (mean difference +3·2%, *P*=0·0003) from 2-3 months to 6 months (**Figure 1**). At 6 months, RVEF tended to be lower in patients with severe illness (58·5±5·1% vs 62·1±6·9%, *P*=0·055).

**Figure 1.**
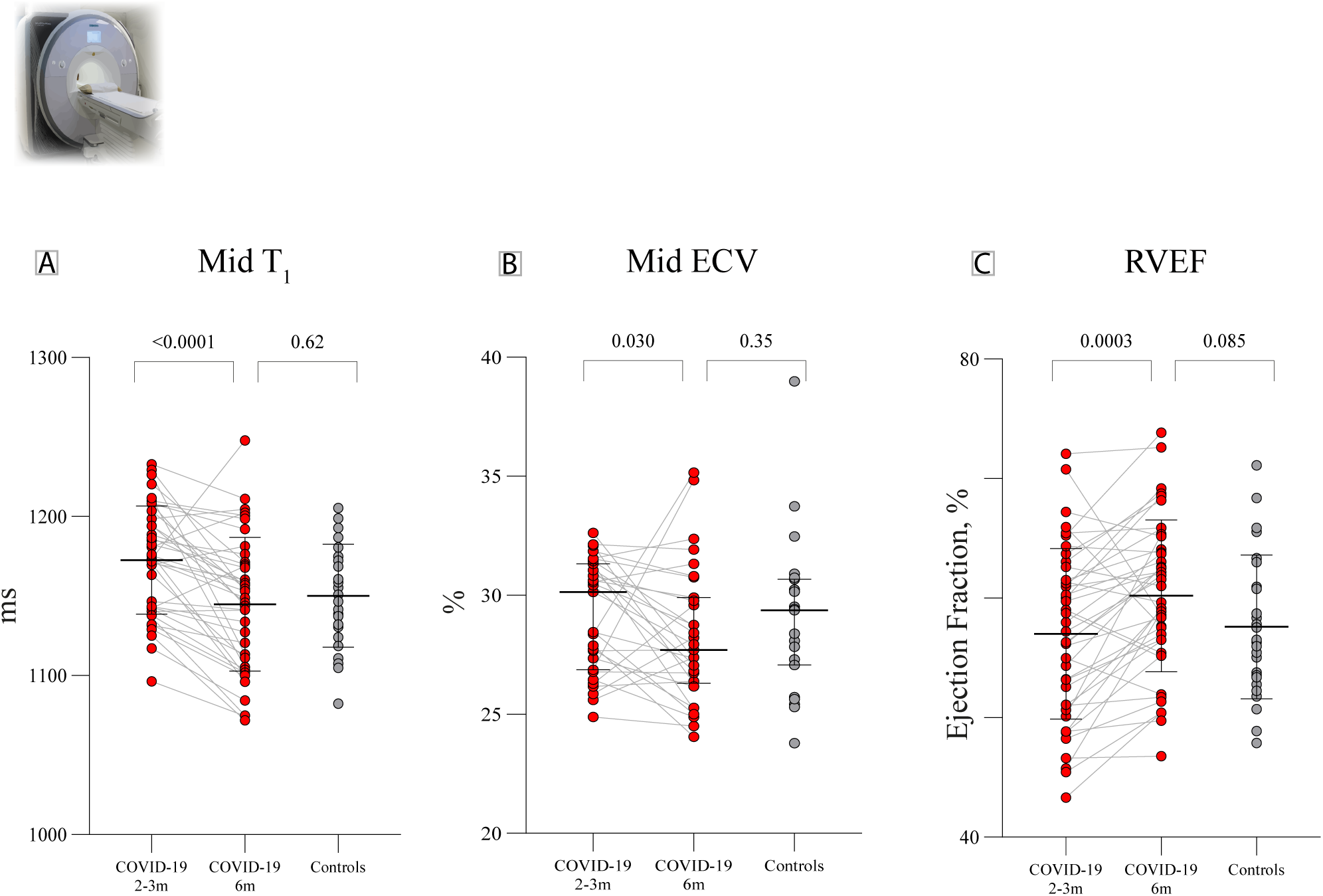
Serial CMR findings in previously hospitalised COVID-19 patients and controls. A: Mid ventricular native T_1_ in patients at 2-3 months was higher than controls, and normalized by 6 months. B: Mid ventricular extracellular volume fraction (ECV) in patients at 2-3 months was comparable to controls, but decreased in patients by 6 months. C: Right ventricular ejection fraction in patients at 2-3 months was comparable to controls, and increased by 6 months. P-values are for group differences (COVID-19 2-3 months vs COVID-19 6 months and COVID-19 6 months vs controls).

Basal and mid-ventricular native T_1_ (a biomarker sensitive to inflammation) values were higher in patients than controls (**Table 4**). By 6 months, myocardial native T_1_ decreased and was no longer different from control T_1_ (**Table 4**; **Figure 1**).

Native T_2_ (a biomarker sensitive to oedema) was not significantly different between patients and controls.

Extracellular volume fraction (ECV, a biomarker sensitive to diffuse fibrosis) did not differ between patients and controls. In patients, slice-averaged ECV decreased (mean difference - 1·13%, *P*=0·005) from 2-3 months to 6 months post-infection.

LGE (measured as % of myocardial volume, a biomarker of focal fibrosis) was slightly higher in patients than controls at 2-3 months (*P*=0·023). By 6 months, this did not differ from controls (*P*=0·62). There were six patients with LGE in a myocarditis pattern and one with evidence of subendocardial infarction (elevated troponin during admission). None of the patients satisfied the updated Lake Louise criteria^20^ for active myocarditis (increased native T_1_/LGE and increased native T_2_) at 6 months.

### Lung imaging and functional assessment

At 2-3 months, 60% of patients had lung parenchymal abnormalities, becoming less extensive (**Table 4**) with time, but were still more common compared to controls at 6 months (*P*<0·0001), Forty percent of patients had lung parenchymal abnormalities involving more than half the lung at 3 months. This reduced to 9% by six months.

At 2-3 months, patients had lower FEV_1_ and FVC compared to controls but most values remained within the normal range (**Table 5**). At 6 months, FEV_1_ was no longer different from controls (*P*=0·10), whereas FVC remained slightly lower (*P*=0·024). Reduced gas transfer (DL_CO_ <80% predicted) and reduced accessible lung volume (V_A_) were seen in 24 patients (52%). Reduced transfer coefficient for carbon monoxide (K_co_) was present in six patients (13%). Patients with parenchymal abnormalities had lower DLco compared to those without (77% vs 91%, *P*=0·009). DLco was not significantly different in patients with severe illness at admission versus non-severe patients (77·4% vs 84·5%, *P*=0·15).

**Table 5.**
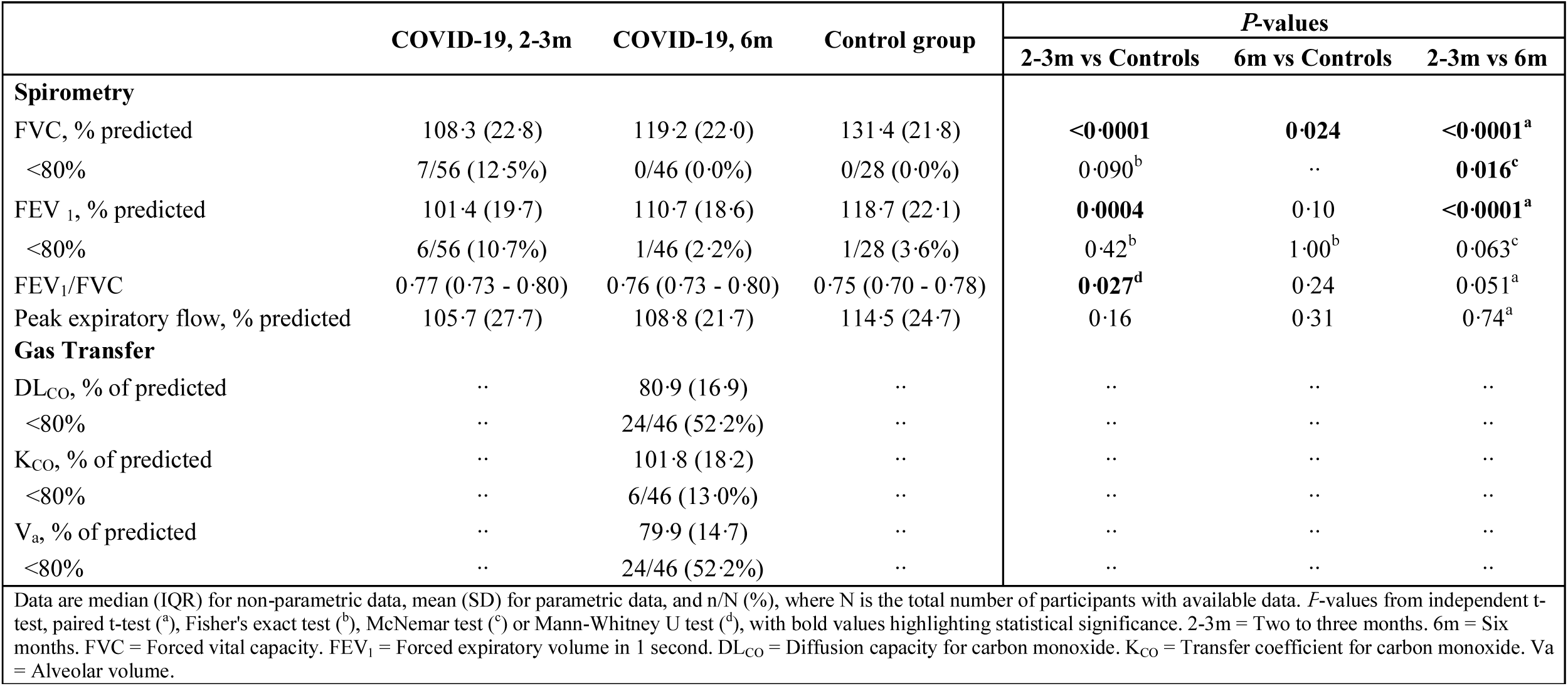
Spirometry and gas transfer testing results in patients at follow-up and controls.

### Serial Cardiopulmonary Exercise Testing

As previously reported, patients had reduced peak oxygen consumption (VO_2_) at 2-3 months. By 6 months, this improved but was still reduced relative to controls (**Table 6, Figure 2**).

**Table 6.**
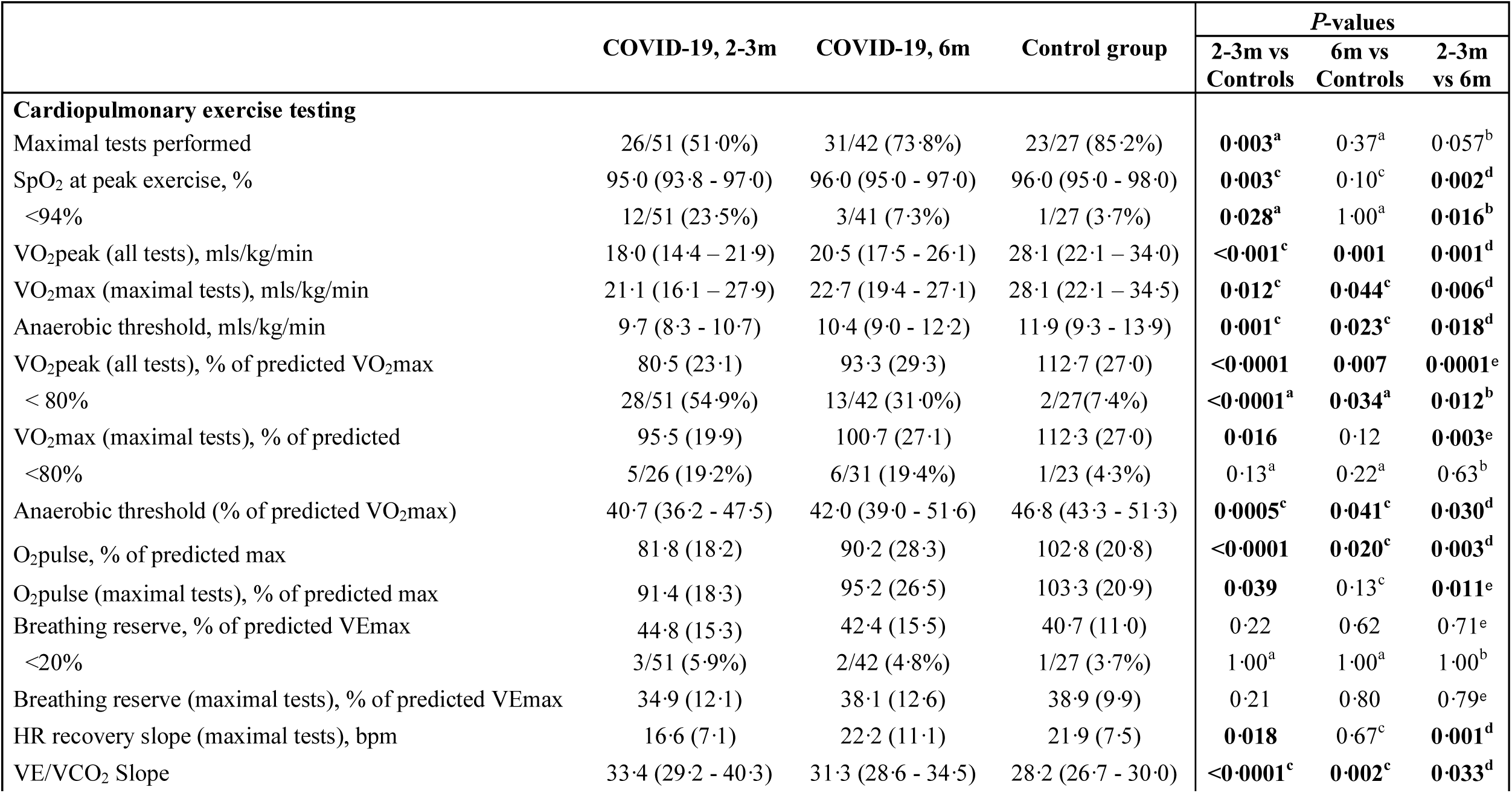
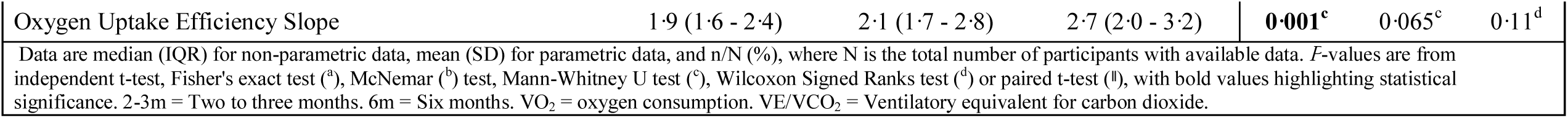
CPET parameters in patients at follow-up and controls.

**Figure 2.**
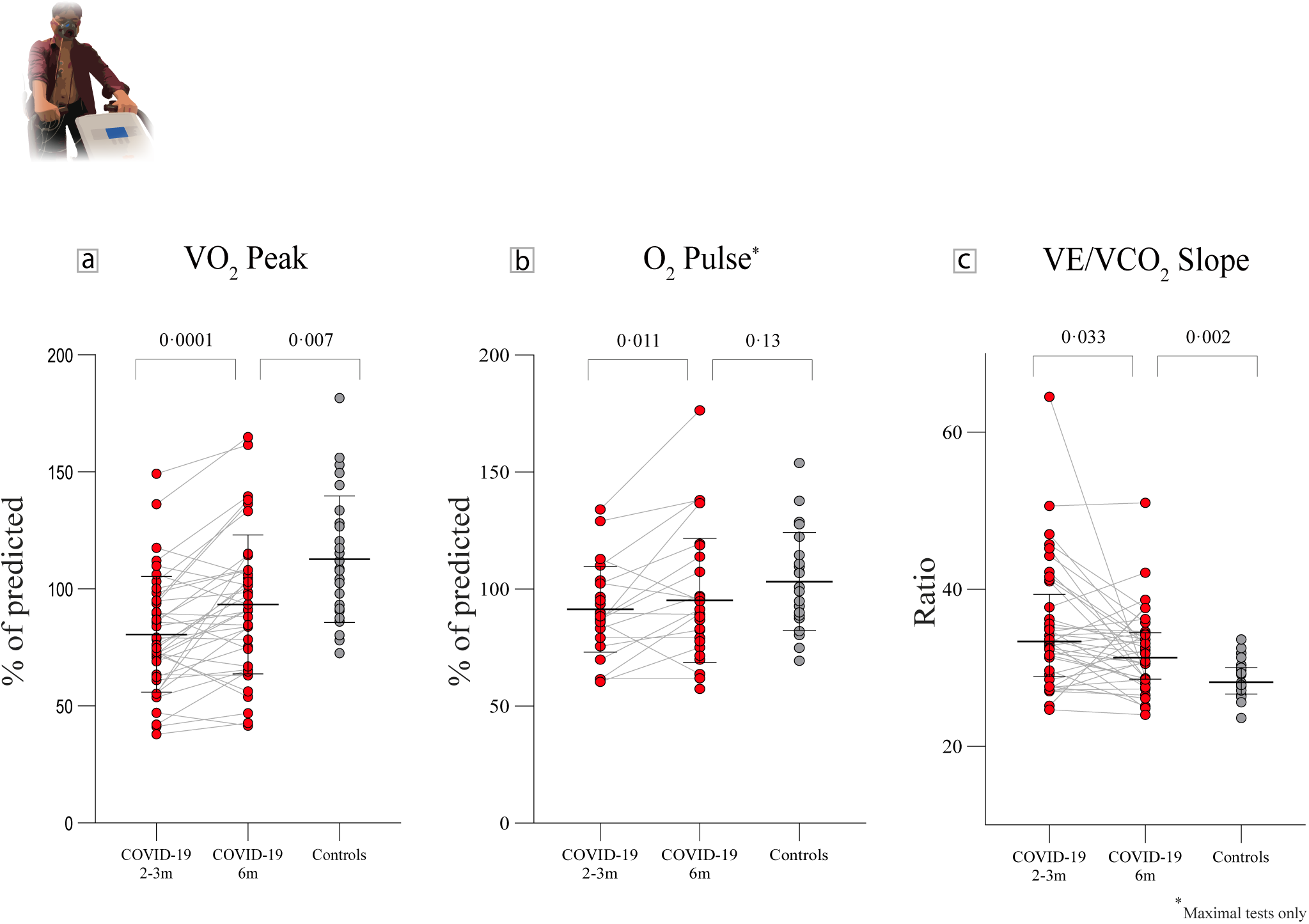
Serial CPET assessments in previously hospitalised COVID-19 patients and controls. A: Peak oxygen consumption (VO_2_ peak) in patients improved from 2-3 months to 6 months, but remained lower than controls. B: Peak oxygen pulse (O_2_ pulse) in patients with maximal tests at 2-3 months was lower compared to controls. By 6 months, this improved and became comparable to controls. C: The ventilatory equivalent for carbon dioxide (VE/VCO_2_) slope in patients improved from 2-3 months to 6 months, but remained high versus controls. P-values are for group differences (COVID-19 2-3 months vs COVID-19 6 months and COVID-19 6 months vs controls).

Maximal test criteria consisted of a respiratory exchange ratio ≥ 1·1 and plateau in oxygen uptake.^21^ At 2-3 months, 49% of patients had submaximal tests (versus 15% of controls, *P*=0·003). By 6 months, this prevalence reduced to 26% (*P*=0·37 for comparison with controls).

In those with a maximal test, maximal VO_2_ was lower in patients at 2-3 months but was no longer so by 6 months (*P*=0·12 for comparison with controls).

The ventilatory equivalent for carbon dioxide (VE/VCO_2_) slope, a marker of ventilatory efficiency, was abnormal in patients at 2-3 months and improved by 6 months (*P*=0·033). In spite of this, the VE/VCO_2_ slope remained borderline abnormal (median 31·3 (IQR 28·6-34·5)) versus controls (median 28·2 (IQR 26·7-30·0, *P*=0·002)). Reduced ventilatory efficiency had little effect on exercise capacity, with respiratory limitation (defined as a breathing reserve of less than 20% at peak exertion) only occurring in 6% and 5% of patients at 2-3 and 6 months, respectively. This did not differ from controls (4%, *P*=1·0).

At 2-3 months, oxygen (O_2_) pulse in maximal tests (a surrogate measure of exercise stroke volume) was lower in patients versus controls and was accompanied by earlier attainment of the anaerobic threshold (AT). By 6 months, O_2_ pulse improved and became comparable to controls (95% of predicted vs 103% of predicted, *P*=0·13). Despite improvement in the AT, occurring later during exercise, it remained different from controls (42% of predicted VO_2_max vs 47% of predicted VO_2_max, *P*=0·041, **Table 6**).

Heart rate recovery (HRR) in the first minute following exercise cessation was slower in patients compared to controls (16·6 vs 21·9 beats, *P*=0·018). By 6 months, HRR improved significantly (22·2 beats, *P*=0·001), and became comparable to controls (*P*=0·67). The severity of illness during admission was not associated with a reduction in peak or maximal oxygen consumption at 2-3 months and 6 months (*P*>0·20 for all comparisons).

### Relationship between symptoms and cardiopulmonary health

At 6 months from infection, neither CMR nor pulmonary function or CPET parameters associated with cardiopulmonary symptoms (**Figure 3**) or breathlessness (**Supplementary Material, p8**). Longitudinal improvement in CMR and CPET parameters did not associate with improvement in cardiopulmonary symptoms from 2-3 months to 6 months (*P*>0·05). There was no correlation between the extent of lung abnormalities on MRI, lung function parameters (FEV_1_, FVC, FEV_1_/FVC, DLco) and breathlessness scores (**Supplementary Material, p5**). The dissociation between physiological measurements and symptoms were further highlighted by the fact that of the twenty patients who did not report significant breathlessness (MRC grade <2) at 6 months, 55% had abnormal gas transfer (DLco <80% predicted).

**Figure 3.**
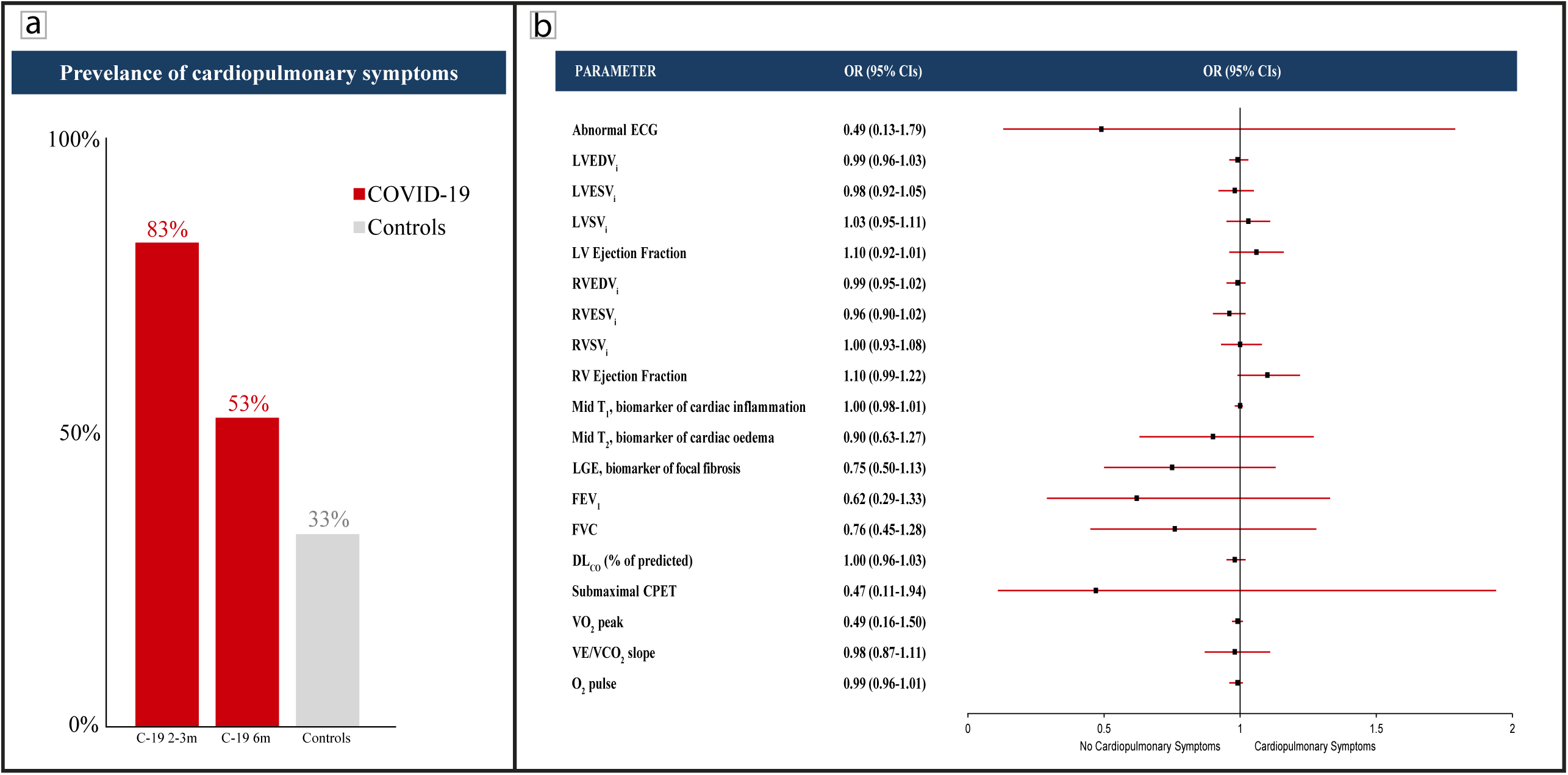
Prevalence and CMR/PFT/CPET determinants of cardiopulmonary symptoms (chest pain, palpitations, syncope, dyspnoea, or dizziness). A: At 2-3 months, 83% of patients experienced at least one cardiopulmonary symptom. By 6 months, this improved to 53% and was comparable to controls. B: Forest plot depicts the odds of having any cardiopulmonary symptom at 6 months using any of the ECG, CMR, PFT, and CPET measures. An abnormal ECG was defined as rhythm abnor-malities and/or the presence of bundle branch block, ST-segment elevation/depression or T wave inversion. CMR - Cardiac magnetic resonance. CPET - Cardiopul-monary exercise testing. OR - Odds ratio. CI - Confidence interval. ECG – Electrocardiogram. LVEDV_i_ - Left ventricular end-diastolic volume (indexed). LVESV_i_ - Left ventricular end-systolic volume (indexed). LVSV_i_ - Left ventricular stroke volume (indexed). RVEDV_i_ - Right ventricular end-diastolic volume (indexed). RVESV_i_ - Right ventricular end-systolic volume (indexed). RVSV_i_ - Right ventricular stroke volume (indexed). LGE - Late gadolinium enhancement. DL_CO_ - Diffusing capacity for carbon monoxide. pVO_2_ - Peak oxygen consumption. VE/VCO_2_ - Ventilatory equivalent for carbon dioxide. O_2_ pulse - Oxygen pulse.

## Discussion

The main findings from our study are as follows: First, serial measures of cardiopulmonary health on CMR in moderate to severe COVID-19 improve over time. Second, exercise tolerance in patients recovers at 6 months post-infection but is still abnormal in some when compared to controls, due to muscular fatigue and weakness. Third, although cardiopulmonary symptom burden improves, more than half the patients remain symptomatic, and neither CMR nor pulmonary function or CPET measures associate with persistent symptom burden.

Since the start of the pandemic, several studies have harnessed the power of CMR to better understand the mechanisms underlying myocardial injury associated with COVID-19.^6,22^ Prevalence estimates of injury have varied due to differences in cohort characteristics and methodologies used. In the largest CMR follow-up study of patients with elevated troponin, Kotecha and colleagues observed that up to 49% of patients have evidence of either myocarditis or myocardial ischemia/infarction .^22^ By contrast, similar-sized studies of younger athletes^23^ and older individuals^6^ with milder infections (predominantly non-hospitalised) have reported variable estimates of myocardial injury (ranging from 1·5% to 70%). The present study is unique to others in the literature, as we prospectively recruited hospitalised COVID-19 patients and risk factor matched controls (who served as our reference) and longitudinally evaluated changes in CMR myocardial tissue characteristics in patients. Here, we show that whilst there were some patients with abnormal myocardial native T_1_ (a marker of oedema and inflammation) at 2-3 months, native T_1_ normalized in the majority by 6 months and was accompanied by a decrease in extracellular volume. These findings highlight two important points. The first is that early tissue abnormalities on CMR are likely due to dynamic alterations in the extracellular environment (hyperaemia^24^ or changes in extracellular proteins/matrix) influenced by circulating cytokines and importantly, not explained by comorbidities alone. This is in line with recent studies that have also demonstrated temporal changes (improvement) in inflammatory cytokines (IL-1, IL-2, IL-6, IL-18, TNF, IFNL1) in COVID-19 patients on serial assessments.^25,26^ The second is that cardiac health is restored in the majority of patients by 6 months. Only two patients had borderline low LV function, RV parameters were normal and there were no cases of active myocarditis (as per the updated Lake Louise criteria^20^). These findings are in keeping with the low prevalence (7%) of cardiac dysfunction (defined by levels of NT-BNP) reported by a large UK-wide prospective follow-up study of post-hospitalised COVID-19 patients by Evans and colleagues.^27^

Exercise intolerance is common among patients recovering from coronavirus infections (SARS, MERS, and COVID-19).^7,8,28,29^ We had previously shown that at 2-3 months^8^, CPET revealed a number of abnormalities in patients. By 6 months, many of these parameters improved, though VE/VCO_2_ slope, AT, and frequency of submaximal tests were still abnormal in patients. Of importance, cardiopulmonary limitations were not felt to be the main driver of exercise intolerance by 6 months as reduced peak oxygen consumption was only seen in symptom-limited submaximal tests. Other CPET studies of COVID-19 patients have also demonstrated a high proportion of submaximal tests.^30^ In one study, direct assessment of maximum muscle strength using dominant leg extension independently predicted peak oxygen consumption in patients following COVID-19.^7^ Taken together, these findings strongly suggest that muscular conditioning and fitness are important determinants of exercise tolerance and highlight the role of dedicated rehabilitation in augmenting recovery.

Postural orthostatic tachycardia and other manifestations of dysautonomia have frequently been described among patients post-COVID-19.^31,32^ Here, we showed that at 2-3 months, heart rate recovery, an indirect measure of autonomic health, was impaired in patients compared to controls.^33^ By six months, heart rate recovery improved, implying that dysautonomia may be transient and does spontaneously recover in some patients.

An interesting observation from serial CPET assessments was that O_2_ pulse, a marker of exertional stroke volume, was reduced in patients at 2-3 months and normalized by 6 months. Reduced O_2_ pulse has also been observed in a recent study by Baratto et al.,^34^ where impaired tissue oxygen extraction has been implicated. Increased pulmonary vascular resistance (PVR) can also explain the reduced O_2_ pulse and may occur secondary to SARS-CoV-2 associated endothelial cell injury, impaired vasodilation,^35^ and/or persistent thrombo-embolic manifestations.^36^ Given that the RV is extremely sensitive to changes in PVR, the observed improvement in O_2_ pulse and RV function by 6 months could reflect restoration of pulmonary vascular homeostasis in patients.

As the COVID-19 pandemic has progressed, our understanding of the long-term effects of SARS-CoV-2 infection has evolved.^37-39^ Multiple studies^5,27^ have demonstrated that some patients recovering from COVID-19 experience a diverse range of persistent symptoms months beyond infection, commonly referred to as “long haul COVID” or “post-COVID-19 syndrome”.^39,40^ In the present study, 1 in 2 patients reported persistent cardiopulmonary symptoms ((chest pain, palpitations, syncope, dyspnoea or dizziness)) at 6 months, despite an improvement in symptoms from 3 months. Neither CMR nor CPET or pulmonary function measures were associated with enduring symptoms. These findings highlight the reduced yield of standard clinical investigations in elucidating a cause for persistent symptoms and the need to explore other mechanisms (sarcopenia, muscle weakness, neurohormonal factors, autoantibodies, nociceptive alterations, mast cell activation syndrome) that may be relevant.^41-45^ Another important finding from this study is that more than half the patients who were asymptomatic had impaired DLco at 6 months, implying that physiological recovery may not be reliably captured by subjective measures of cardiopulmonary health. Further efforts are needed to better understand the determinants of impaired DLco and persistent parenchymal abnormalities associated with COVID-19, as we seek to develop effective treatments that could potentially reverse the long-term sequelae of COVID-19.

Although the sample size of this study is small, it has many strengths. To our knowledge, this is the first study to comprehensively (cardiopulmonary imaging, static physiology, whole-body exercise testing, patient health questionnaires) evaluate the longitudinal trajectory of cardiopulmonary abnormalities on CMR and CPET in patients at 3 and 6 months. From a diagnostic perspective, our study provides important insight into the lack of association between symptoms and results from standard clinical investigations. The longitudinal design and incorporation of risk factor matched control group clarified the relevance of some early abnormalities. While it was difficult to exclude significant inducible ischemia, none of the patients stopped their CPET because of angina or ischemic ECG changes. Arterial blood gas sampling or echocardiography during CPET were lacking and did not permit assessment of tissue oxygen extraction, cardiac output during exercise and pulmonary dead space. Not all patients came back for follow-up assessments (due to work commitments or had moved abroad; see supplement for details) which could in turn bias prevalence estimates in this study. However, this would not affect the relationship between symptoms and objective measures of cardiopulmonary health.

## Conclusion

Our study provides novel insights into the trajectory of cardiopulmonary symptoms and abnormalities on serial CMR, spirometry and CPET in patients. At 6 months, cardiac abnormalities on CMR improved in the majority of patients and were not different to matched controls. Parenchymal abnormalities, lung function impairment and CPET improved but were still abnormal relative to controls. Nearly half the patients continue to experience symptoms at 6 months. There was a surprising dissociation between persistent cardiopulmonary symptoms and CMR/CPET parameters, underscoring the need to examine alternative mechanisms for symptom persistence in patients.

## Data Availability

The data underlying this article will be shared on reasonable request to the corresponding author, subject to institutional and ethical committee approvals.

## Acknowledgements

The authors’ work was supported by the NIHR Oxford and Oxford Health Biomedical Research Centre, Oxford British Heart Foundation (BHF) Centre of Research Excellence, United Kingdom Research Innovation and Wellcome Trust. This project is part of a tier 3 study (C-MORE) within the collaborative research programme entitled PHOSP-COVID Post-hospitalization COVID-19 study: a national consortium to understand and improve long-term health outcomes. Funded by the Medical Research Council and Department of Health and Social Care/National Institute for Health Research Grant (MR/V027859/1) ISRCTN number 10980107.

This work also arises from one of the national “COVID-19 Cardiovascular Disease Flagship Projects” designated by the NIHR-BHF Cardiovascular Partnership.

The sponsors played no role in the design of the study; collection, analysis and interpretation of data; in writing the manuscript, and in the decision to submit the paper for publication. The views expressed are those of the authors and not necessarily those of the National Health Service, NIHR, or the United Kingdom Department of Health.

All authors had full access to all the data in the study and accept responsibility to submit for publication.

We thank our participants and their families who have given their time to help others understand the medium to long-term effects of COVID-19. We are grateful to the University of Oxford and Oxford University Hospital Trust for their support of this study. We would like to acknowledge OCMR staff, Ms Hanan Lamlum, Ms Rebecca Mills, Ms Polly Whitworth, Ms Claudia Nunes, Ms Harriet Nixon, Ms Liliana Da Silva Rodrigues, Ms Kinga Varnai and Ms Catherine Krasopoulos for their help with this work. We acknowledge the support of Siemens in providing WIP 1048 for cardiac T_1_ mapping.

