## Supplementary Material for "Symptom Persistence Despite Improvement in Cardiopulmonary Health – Insights from longitudinal CMR, CPET and lung function testing post-COVID-19"

**Methods**

**Inclusion Criteria**

We included all patients with moderate to severe acute respiratory distress syndrome Coronavirus-2 (SARS-CoV-2 infection) (see below for definition), confirmed by a reverse transcription and polymerase chain reaction (RT-PCR) nasopharyngeal swab test.

Controls were non-hospitalized subjects (invited from the community) without symptoms or signs of a respiratory tract infection of coronavirus disease, who were screened for SARS-CoV-2 antibodies and tested negative.

**Exclusion Criteria**

Subjects with contraindications to magnetic resonance imaging (metal implant in body, known claustrophobia, pacemakers, contrast allergy) and severe comorbidities – end-stage renal, cardiac, liver, neurological disease – were excluded from the study.

**Definitions**

Moderate to severe SARS-CoV-2 infection: Severity of acute illness on admission was defined by the World Health Organisation (WHO) interim clinical guidance. All patients with clinical signs of pneumonia such as respiratory rate > 30 breaths/min; or severe respiratory distress; or SpO2 < 90% (on room air) and requiring hospital admission for more than 48 hours were assessed to have moderate to severe coronavirus disease (COVID-19).[1]

Severity of disease (or clinical response) during admission: Patient response in hospital was defined by the WHO ordinal scale for clinical improvement.[2] A score 0 was given to an uninfected individual, 1 to an ambulatory patient without limitations of activities, 2 where there were limitations of activities, 3 for a hospitalized patient not on oxygen ($O_2$) therapy, 4 for $O_2$ supplementation via simple face mask or nasal prongs, 5 for the administration of non-invasive ventilation (NIV) or high flow $O_2$, 6 for being intubated and on mechanical ventilation, 7 for requiring ventilation and additional organ support such as renal replacement therapy and extracorporeal membrane oxygenation (ECMO), and 8 for death.[2]

Lymphocytopenia**:** Defined as a lymphocyte count of less than 1000 cells per cubic millimetre.[3]

Thrombocytopenia: Defined as a platelet count of less than 100,000 per cubic millimetre.[3]

Acute kidney injury: Acute kidney injury (AKI) was defined based on the highest serum creatinine level and urine output. A diagnosis of AKI was made based on any of the following: an increase in serum creatinine levels by 0.3 mg/dl or greater (26.5 μmol/l or greater) within 48 hours; an increase in serum creatinine levels to 1.5 times of the baseline level or greater, which was known or presumed to have occurred within 7 days; or urine volume of below 0.5 ml/kg/h for 6 consecutive hours.[3]

Acute cardiac injury: Acute cardiac injury was suspected when serum levels of cardiac biomarkers (high-sensitivity cardiac troponin I) were above the $99^{th}$ percentile upper limit of normal or new abnormalities were seen on electrocardiography and echocardiography.[3]

Acute liver injury: Acute liver injury was defined as blood levels of alanine aminotransferase (ALT) or aspartate aminotransferase (AST) above three times the upper reference limit, alkaline phosphatase or gamma-glutamyl transferase levels above two times the upper reference limit.[4]

**Questionnaires**

**Patient health questionnaire (PHQ-15)**

The Patient Health Questionnaire-15 is an instrument used for assessing the burden of somatic symptoms.[5] This scale was used to assess for the presence of a number of symptoms common in patients following COVID-19 infection.

**Medical Research Council scale (MRC)**

The MRC dyspnoea scale[6] was used to assess the effects of breathlessness on daily activities. This scale was used to measure perceived respiratory disability on a scale of 1 (no breathlessness) to 5 (complete incapacity).

**Fatigues Severity Scale**

The Fatigue Severity Scale (FSS) is a symptom-specific outcome scale aimed at providing a standardized means of describing health state.[7] The FSS comprises of nine statements, describing the severity and impact of fatigue, with a scale of possible responses ranging from 1 (“strongly disagree”) to 7 (“strongly agree”). FSS was reported as the mean score over the nine items; a higher mean score (≥4) indicates greater severity.

**MRI**

Scans were carried out during a single scan session on a Siemens Prisma 3T, with a Siemens 32-channel spine array and 30-channel body array used for cardiac and pulmonary imaging.

**Lung MRI**

A) Free breathing half-Fourier-acquisition single-shot turbo spin-echo (HASTE) MRI – [Typically axial and coronal; TR/TE = 750/49 ms; R=2; field of view = 380x320 mm,$^2$ slice thickness/spacing 8/0mm, 35 slices, matrix = 256x123]. T2-weighted HASTE images provide high-signal intensity in water-rich tissues. Lung abnormalities tend to appear bright, surrounded by air-filled parenchyma with low signal intensity.

**Lung MRI Analysis**

Axial and coronal thoracic HASTE images were qualitatively read by an expert radiologist (CX) and scored as 0 (0%), 1 (1–25%), 2 (26–50%), 3 (51–75%), or 4 (76–100%) depending on the extent of parenchymal involvement.

**Cardiac MRI**

The cardiac MRI protocol included routine clinical and advanced parametric mapping sequences. The cardiac MRI sequences were as follows:

A) Cine steady-state free precession (SSFP) imaging - [Three long-axis and short-axis stack; Typically TR/TE = 3.16/1.38ms; R = 3; flip angle = 65°; field of view = 360×270 mm, matrix 208×139, slice thickness/spacing 7/3mm]. This is used in the clinical setting to assess cardiac (left and right ventricular) mass, volumes, and function which may be affected in cardiovascular diseases.

B) Shortened Modified Look Locker $T_1$ mapping (ShMOLLI, Siemens prototype sequence WIP1048B) [Base, mid, apex short axis slices; TR/TE = 2.7/1.07ms, R = 2, flip angle = 35°, field

of view = 360×270 mm, matrix = 192×144; slice thickness = 8 mm. Inversion times in image acquisitions, waiting periods with conditional on-the-scanner $T_1$ map reconstruction as described previously] [8]. This sequence helps assess the myocardial tissue characteristics which indicates if the myocardium has increased water content or fibrosis.[9]
C) $T_2$ mapping (Siemens Myomaps) - [Base, mid, apex short axis slices; bSSFP readout; TR/TE = 3/1.3ms, R = 2, flip angle = 20°, Matrix = 192×142, field of view = 360×270 mm, slice thickness = 8 mm]. This sequence is typically used to detect increased water content or swelling in myocardial tissue.
D) Late gadolinium imaging was acquired using a $T_1$-weighted phase-sensitive inversion recovery sequence following a bolus injection of 0.15 mmol/kg of body weight of gadolinium-based contrast agent (Dotarem) and a 10 ml saline flush. Typical scan parameters: [TR/TE/TI = 2.9/3.36/subject-specific ms; R = 2; flip angle = 55°; matrix = 144×256, field of view 380×285 mm, slice thickness = 8 mm.] Late gadolinium imaging is used in clinical cardiac MRI to detect regions of focal fibrosis or scar within the myocardium.
E) $T_1$-weighted imaging: Base, mid, and apical $T_1$ maps (ShMOLLI) were acquired 15 minutes after the administration of contrast. This sequence is used to estimate the extent of diffuse fibrosis or scar in the myocardium.

**Cardiac MRI image analysis**

All cardiac MRI images were analysed using cvi42 software (Circle Cardiovascular Imaging Inc., Version 5.11.4, Calgary, Canada). Blinded analyses of cardiac images were undertaken by BR, MC. Volume and function: Left and right ventricular short-axis epicardial and endocardial borders were manually contoured at end-diastole and end-systole (endocardial border only). Left and right ventricular end-diastolic (EDV) and end-systolic (ESV) volumes were used to calculate stroke volume (SV) as SV = EDV-ESV.[10] Ejection fraction (EF) was calculated as a ratio of SV and EDV (EF = SV/EDV). LV mass was calculated by subtracting the endocardial volume from the epicardial volume, based on prior knowledge of myocardial specific gravity (1.05 g/cm$^3$).[11] Volumes and mass were indexed to body surface area.
Analysis of $T_1$ and $T_2$ maps: All (pre and post-contrast) $T_1$ maps underwent strict quality control as previously described [9] by an observer who underwent standardized institutional training.[12,13] $R^2$ maps of ShMOLLI inversion recovery fit were used to verify the quality of $T_1$-maps on the scanner. Manual epicardial and endocardial contours were drawn on base, mid and apical slices, and average slice $T_1$ was derived. Extra-cellular volume (ECV) for the three slices was calculated from pre-and post-contrast $T_1$ maps and haematocrit (HCT) using the formula: $ECV = (\Delta[1/T_1 myocardium]/\Delta[1/T_1 blood]) * [1-Hct])$.
All $T_2$ maps also underwent strict quality control. Manual epicardial and endocardial contours were drawn on base, mid and apical slices (acquired at identical slice positions to pre and post-contrast $T_1$ maps). Average slice $T_2$ was derived for each slice.[14]
Late gadolinium enhancement (LGE): Endocardial and epicardial borders were drawn on the phase-sensitive inversion recovery (PSIR) images alone with a reference region of interest (ROI) with normal myocardial intensity as 'remote' region. Hyperenhanced pixels were defined as those that were five standard deviations (5-SD) above the mean intensity of the reference ROI placed in a remote area of myocardium with no visual evidence of enhancement.[14]
Blinded qualitative assessment of LGE images was undertaken by an experienced CMR reader (BR) and visually classified as the following: 1) no LGE, 2) myocarditis pattern, 3) myocardial infarction, 4) LV/RV insertion point fibrosis, 5) mixed (infarction and myocarditis), 6) other (cardiomyopathy). All cases were also assessed for presence of pericardial effusion.[15]

**Spirometry and Gas transfer**

At least three spirometry measurements were performed, until acceptability criteria defined by the American Thoracic Society (ATS) and European Respiratory Society (ERS) were met.[16] Bronchodilators were not administered. The largest $FEV_1$ and FVC of the triplicate readings and respective $FEV_1$/FVC ratio were reported.

Gas transfer assessment was carried out using a ten-second single breath hold technique, with methane as the tracer gas. Assessment was repeated until acceptability and repeatability criteria, as defined by ATS/ERS, were met.[17] The measurements acquired (diffusion capacity for carbon monoxide (DLco) and alveolar volume (Va) were adjusted for haemoglobin and standard pressure (760mm Hg).

**Cardiopulmonary exercise test (CPET)**

All patients performed symptom-limited incremental cycle ergometer exercise testing with electrocardiographic monitoring. The gas analyser was calibrated prior to each test. Each subject was fitted with a facemask incorporating a disposable flow turbine.

Continuous 12-lead electrocardiographic recordings were acquired for each subject. Oxygen saturation via pulse oximetry was recorded continuously. Blood pressure was measured with a manual sphygmomanometer every 3 to 4 minutes. Following 2 minutes of unloaded cycling, the work rate was increased to 20W. This was followed by a 10W/min ramp. Predetermined conditions for termination of the test included arrhythmia, chest pain, hypotensive response during exercise, or an excessive hypertensive response during the test (>250/115 mm Hg). Otherwise, participants were encouraged to reach their maximal work rate. Following peak effort, participants performed a further 3 minutes of unloaded cycling.

Maximal voluntary ventilation (MVV) was calculated by multiplying $FEV_1$ by 40. Peak oxygen uptake ($VO_2$peak) was predicted using the Wasserman weight algorithm.[18] Maximum heart rate was predicted using the equation 220 – age in years.[19] The V-slope method was used to identify the anaerobic threshold (AT). This was normalized to peak $VO_2$ as ($VO_2$ at AT/predicted peak $VO_2$) x 100). All data points were averaged every 20 seconds.

**Impact of ethnicity on exercise parameters and relationship between objective measures of health**

To assess the impact of ethnicity on differences in exercise parameters at 6 months, two general univariate linear models were used, one with oxygen consumption at peak exercise as the dependent variable and the other with $\dot{V}E/\dot{V}CO_2$ slope as the dependent variable. Participant status (COVID-19 patient or control) was used as a fixed factor, and BAME status was used as a covariate. Differences in oxygen consumption at peak exercise and $\dot{V}E/\dot{V}CO_2$ between COVID-19 patients and controls persisted when correcting for ethnicity (peak oxygen consumption p-value 0.031 and $\dot{V}E/\dot{V}CO_2$ p-value 0.0004).

To assess the impact of ethnicity on the relationship between CMR/CPET parameters and persistent cardiopulmonary symptoms at 6 months, binary logistic regression was performed as described in the manuscript, with the addition of BAME status for each independent variable assessed. The absence of a significant relationship between CMR/CPET parameters and persistent cardiopulmonary symptoms at 6 months persisted.

**Correlation between breathlessness, pulmonary function and MRI parenchymal abnormalities**

No significant correlations were observed between breathless (defined as MRC dyspnoea scale >1) and $FEV_1$ (rho=0.036), FVC (rho=0.020), FEV1/FVC (rho=-0.261), DLco (rho=0.010) and the extent of lung parenchymal abnormalities on MRI (rho=-0.004) (p>0.05 for all assessments).

**Table 1. Details on missing data for variables in COVID-19 patients at 6 months and controls.**

| Cohort, variable/s | Data, frequency (%) | Reason |
|---|---|---|
| **Lung and Cardiac imaging** | | |
| COVID-19, all variables | 44/46 (95.7%) | 2 did not undergo imaging due to claustrophobia |
| COVID-19, late gadolinium enhancement and post-contrast $T_1$ | 43/46 (93.5%) | 2 did not undergo imaging due to claustrophobia, and 1 opted not to received gadolinium |
| CONTROL, all variables | 28/30 (93·3%) | 2 did not undergo imaging due to claustrophobia |
| CONTROL, Post-contrast $T_1$ | 26/30 (86.6%) | 2 did not undergo imaging due to claustrophobia, 2 failed QC |
| CONTROL, all variables | 28/30 (93·3%) | 2 did not undergo imaging due to claustrophobia |
| **Cardiopulmonary exercise testing** | | |
| COVID-19, all variables | 42/46 (91.3%) | 4 were unable to perform the test (1 due to large body habitus, 1 due to claustrophobia on wearing a mask, 1 due to acrophobia when sitting on the bicycle, and 1 due to work commitments limiting the study visit) |
| CONTROL, all variables | 27/30 (90·0%) | 1 was unable to perform the test (due to large body habitus), 2 did not consent to the test |

Data are n/N (%), where N is the total number of participants with available data.

**Figure 1. Recruitment flow-chart of A) patients and B) Controls. *see supplementary material Table 1 for details.**

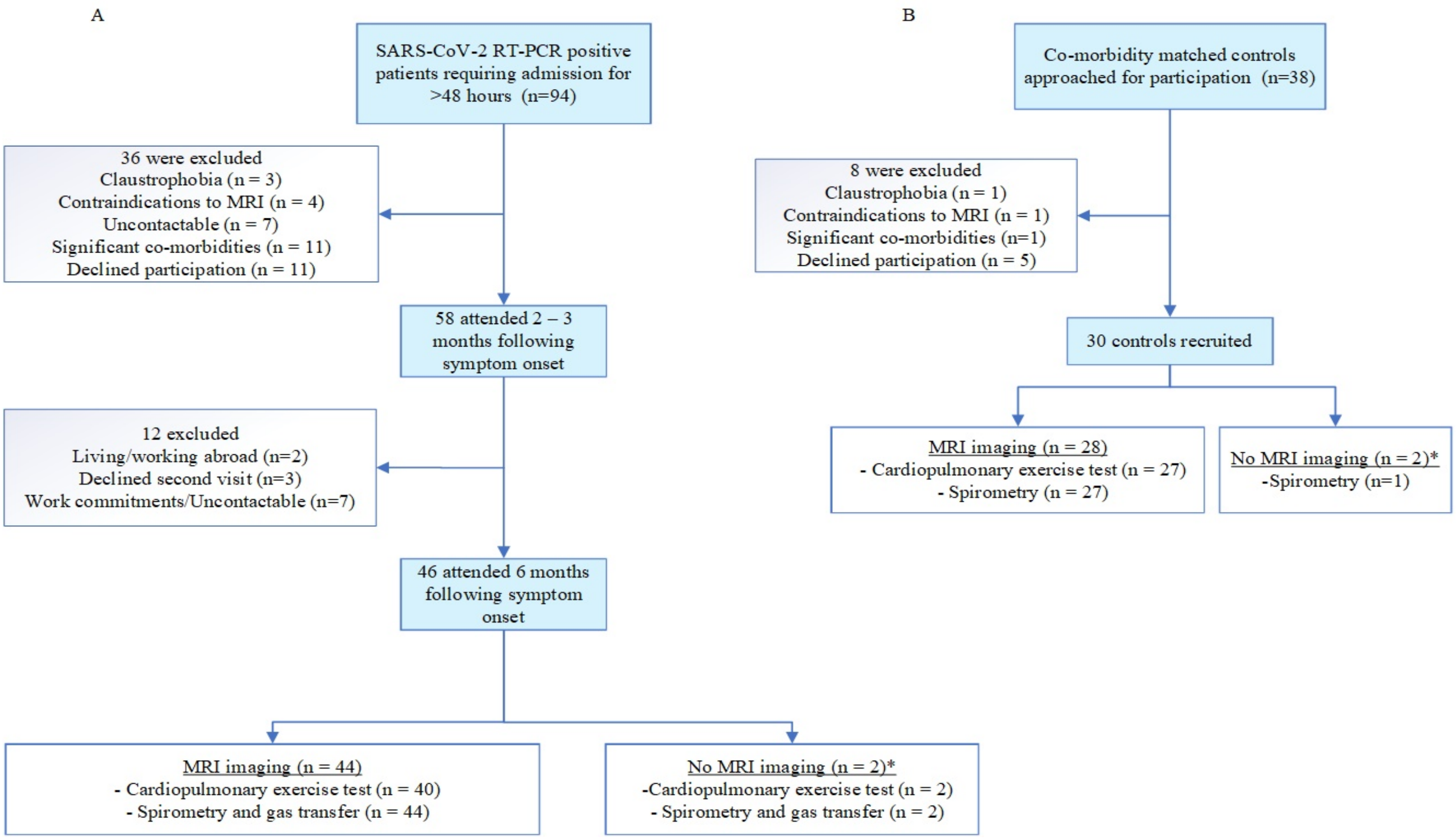

**Figure 2. CMR/PFT/CPET determinants of shortness of breath (MRC dyspnoea scale >1).**

| PARAMETER | OR (95% CIs) |
|---|---|
| Abnormal ECG | 1.16 (0.32-4.21) |
| LVEDVi | 0.99 (0.96-1.03) |
| LVESVi | 0.99 (0.92-1.05) |
| LVSVi | 0.98 (0.91-1.06) |
| LV Ejection Fraction | 1.10 (0.92-1.01) |
| RVEDVi | 0.99 (0.95-1.03) |
| RVESVi | 0.97 (0.91-1.03) |
| RVSVi | 1.01 (0.94-1.09) |
| RV Ejection Fraction | 1.05 (0.95-1.16) |
| Mid $T_1$, biomarker of cardiac inflammation | 1.00 (0.98-1.01) |
| Mid $T_2$, biomarker of cardiac oedema | 0.95 (0.68-1.35) |
| LGE, % myocardial enhancement | 1.34 (0.75-2.39) |
| $FEV_1$ | 0.99 (0.96-1.02) |
| FVC | 1.00 (0.97-1.03) |
| $DL_{CO}$ (% of predicted) | 1.00 (0.96-1.03) |
| Submaximal CPET | 0.69 (0.17-2.73) |
| $VO_2$ peak | 0.49 (0.16-1.50) |
| $VE/VCO_2$ slope | 1.11 (0.96-1.28) |
| $O_2$ pulse | 0.98 (0.95-1.01) |

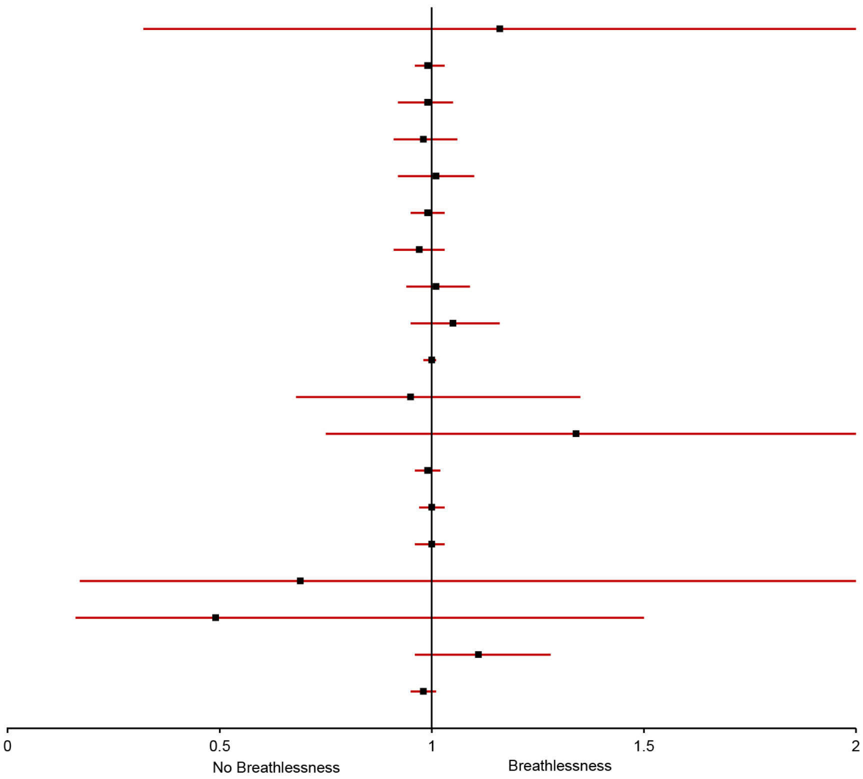

Forest plot depicting outcome analysis (univariate binary logistic regression) with breathlessness (Medical Research Council dyspnoea scale >1) as the outcome variable, demonstrating that CMR, PFT and CPET parameters did not predict breathlessness in COVID-19 patients 6 months following disease onset. Abnormal ECG defined as rhythm abnormalities and/or presence of bundle branch block, ST elevation/depression or T wave inversion. CMR – Cardiac magnetic resonance. PFT – Pulmonary function test. CPET – Cardiopulmonary exercise test. OR - Odds ratio. CI - Confidence interval. ECG – Electrocardiogram. LVEDVi - Left ventricular end-diastolic volume (indexed). LVESVi - Left ventricular end-systolic volume (indexed). LVSVi - Left ventricular stroke volume (indexed). LV – Left ventricle. RVEDVi - Right ventricular end-diastolic volume (indexed). RVESVi - Right ventricular end-systolic volume (indexed). RVSVi - Right ventricular stroke volume (indexed). RV – Right ventricle. LGE - Late gadolinium enhancement. $FEV_1$ – Forced expiratory volume in the first second. FVC - Forced vital capacity. DLco - Diffusing capacity for carbon monoxide. $pVO_2$ - Peak oxygen consumption. $VE/VCO_2$ - Ventilatory equivalent for carbon dioxide. $O_2$ pulse - Oxygen pulse.)
